# Gender-associated differences in serum metabolite profiles among patients with Parkinson’s Disease

**DOI:** 10.1101/2025.10.30.25339159

**Authors:** Carmen Marino, Federica Carrillo, Tommaso Nuzzo, Matteo Vidali, Marcello Serra, Sara Pietracupa, Nicola Modugno, Manuela Grimaldi, Anna Maria D’Ursi, Francesco Errico, Teresa Esposito, Alessandro Usiello

**Affiliations:** Department of Pharmacy, University of Salerno, 84084, Fisciano, Salerno, Italy; Institute of Genetics and Biophysics, Italian National Research Council CNR, 80131, Naples, Italy; Department of Environmental, Biological and Pharmaceutical Sciences and Technologies, Università degli Studi della Campania "Luigi Vanvitelli", 81100, Caserta, Italy; CEINGE Biotecnologie Avanzate Franco Salvatore, Naples, Italy; Clinical Pathology Unit, Fondazione IRCCS Ca’ Granda Ospedale Maggiore Policlinico, Milano, Italy; Department of Biomedical Sciences, University of Cagliari, Monserrato, Italy; IRCCS INM Neuromed, 86077, Pozzilli, Italy; Department of Human Neuroscience, Sapienza University of Rome, Italy; Department of Agricultural Sciences, University of Naples “Federico II”, 80055, Portici, Italy

**Keywords:** serum metabolomics, sex differences, amino acid metabolism, ¹H-NMR, lipid metabolism

## Abstract

Parkinson’s disease (PD) is increasingly recognized as a multisystem disorder associated with peripheral metabolic abnormalities. However, the extent to which sex and genetic background influence circulating metabolomic alterations remains unclear. Using ^1^H-Nuclear Magnetic Resonance (NMR)-based metabolomics, we profiled serum metabolites from 245 clinically and genetically characterized patients with PD and 137 sex-matched healthy controls (HCs). The PD cohort comprised 121 patients with idiopathic PD and 124 patients carrying at least one pathogenic variant in *LRRK2*, *TMEM175*, *PARK2*, *PINK1*, *PARK7*, or *GBA1*.

NMR profiling revealed marked metabolic differences between cases and controls, identifying sex as a major determinant of the circulating metabolic signature. Compared with HCs, male patients exhibited prominent alterations in amino acid metabolism, characterized by reduced glutamate and increased tryptophan levels, together with changes in metabolites related to energy homeostasis. By contrast, female patients displayed a distinct metabolic profile characterized by altered threonine and choline levels, together with perturbations in phosphatidylethanolamine, fatty acids, and ketone bodies.

Notably, despite PD subtype-specific metabolic differences relative to sex-matched controls, multivariate analyses did not discriminate idiopathic from genetic PD in either sex, suggesting that shared disease-related metabolic alterations predominate within the resolution and metabolite coverage of the NMR platform.

Overall, our findings identify biological sex as a major determinant of circulating metabolomic alterations in PD, whereas idiopathic and genetic forms largely converge towards a common systemic metabolic phenotype.

## Introduction

Parkinson’s disease (PD) is a neurodegenerative disorder characterized by the accumulation of alpha-synuclein and the progressive loss of dopaminergic neurons in the substantia nigra pars compacta ^1–3^. Approximately 85–90% of PD cases are classified as idiopathic (iPD), meaning that no highly penetrant causative mutation can be identified ^4^. PD is currently considered a multifactorial disorder arising from the complex interplay between aging, genetic susceptibility, and environmental exposures ^5^. Although patients with iPD do not carry monogenic pathogenic variants, genome-wide association studies have identified multiple risk loci that contribute to disease susceptibility and may influence clinical heterogeneity, age at onset, and disease progression ^6^. Idiopathic PD is associated with mitochondrial dysfunction, oxidative stress, impaired autophagy- lysosomal pathways, neuroinflammation, and systemic metabolic alterations, suggesting that peripheral metabolic changes may mirror central neurodegenerative processes ^7–15^.

Approximately 10-15% of PD cases are associated with pathogenic variants in genes involved in disease development. Previous studies have shown that genetic PD (gPD) is highly heterogeneous, with pathogenic variants occurring in multiple genes, each identified in only a small proportion of patients. The most frequently implicated genes include *GBA1*, *LRRK2*, *PARK2*, *PARK7*, *PINK1*, and *TMEM175*, which contribute to PD pathogenesis and risk through diverse biological mechanisms, including protein misfolding, mitochondrial function, and impaired cellular detoxification^16–19^.

Importantly, whether idiopathic and genetic PD represent clinically indistinguishable entities or partially divergent phenotypes remains an area of active investigation. While the core motor features—bradykinesia, rigidity, and resting tremor—are largely shared across PD subtypes, several studies suggest genotype-specific differences in age at onset, disease progression, non-motor burden, and therapeutic response^20–23^.

Biological gender also plays a crucial role in PD pathophysiology ^24–27^. Men have a higher risk of developing PD than women, with male-to-female incidence ratios generally reported around 1.5 ^28,29^. This sex difference likely reflects the combined influence of hormonal, genetic, epigenetic, immune and metabolic factors, together with differential environmental and lifestyle exposures ^25,27,30^. Sex-related differences have also been reported in symptom profile, non-motor manifestations, disease progression, and treatment response ^31,32^.

Recent advances in metabolomics have greatly improved our understanding of PD pathophysiology, highlighting complex metabolic dysregulation across multiple biofluids that may contribute to the clinical heterogeneity of the disease ^15,33–42^. However, metabolomic studies in PD have yielded partly inconsistent results, likely owing to differences in study design, analytical platforms, biofluid type, disease stage, medication status, and cohort composition. Importantly, most available studies have involved relatively small cohorts and have not systematically accounted for two major sources of biological heterogeneity in PD: genetic background and gender differences ^37^. As a result, it remains unclear to what extent the metabolic heterogeneity reported across studies reflects shared disease-related mechanisms, genetic background or sex-dependent biological differences. Therefore, large, well-characterized cohorts integrating clinical, genetic and gender stratification are needed to disentangle shared and subtype-specific metabolic signatures.

To address this gap, we conducted ^1^H-NMR-based serum metabolomics in a large cohort of clinically and genetically characterized PD patients (n = 245) and sex-matched healthy controls (HCs, n = 137). Patients were classified into iPD (n = 121) and gPD (n = 124), with the latter carrying at least one pathogenic variant in *LRRK2, TMEM175, PARK2, PINK1, PARK7*, or *GBA1*. By integrating metabolomic profiling with genetic and sex stratification, we aimed to identify shared and subtype-specific metabolic signatures associated with PD.

## Results

### Clinical and demographic characteristics of the NMR study cohort

A total of 137 HCs and 245 PD patients were included in this study. Their demographic and clinical features are reported in **Table 1**. Within the PD cohort, 121 patients were classified as idiopathic PD (iPD), as they tested negative for pathogenic mutations or variants in PD-associated genes (65 males, 56 females). The remaining 124 patients carried PD-associated genetic variants in the most frequently mutated genes, including *LRRK2, GBA1, TMEM175, PINK1, PARK2,* and *PARK7*, and were therefore classified as genetic PD (gPD; 64 males, 60 females) **(Table 2)**. HCs ranged 43-90 years, whereas iPD and gPD ranged 48-82 and 41-85 years, respectively. Both the iPD and gPD patients were slightly older than HCs (median age: 68 vs 62 years), whereas sex distribution did not differ across groups **(Table 1)**. The two PD subtypes showed comparable age at onset, levodopa equivalent daily dose (LEDD) and motor severity as assessed by the Movement Disorders Society Unified Parkinson’s Disease Rating Scale, part III (MDS-UPDRS III) (**Table 2)**. A significant difference was observed in disease duration, which was slightly longer in gPD than in iPD patients (median: 6 vs 5 years, respectively) (**Table 2)**.

**Table 1.**
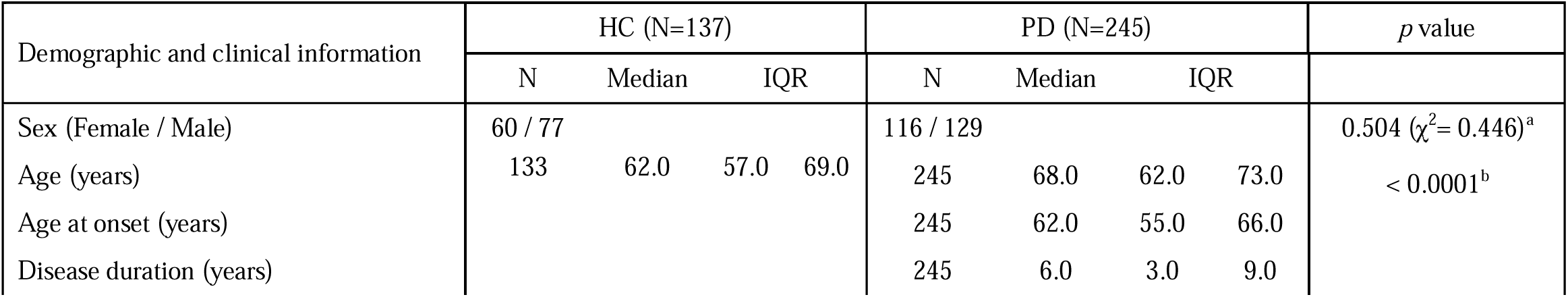
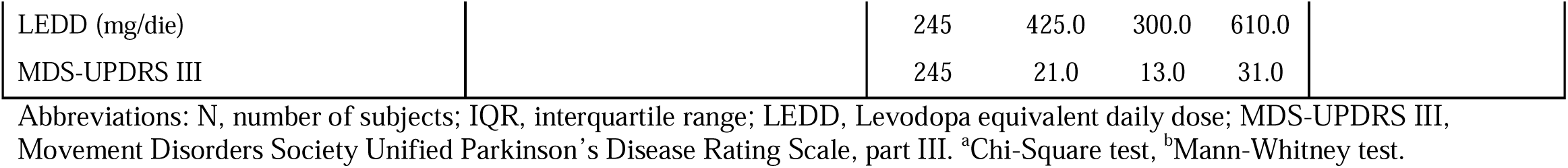
Demographic and clinical characteristics of healthy controls and PD patients enrolled in the serum collection for NMR analysis.

**Table 2.**
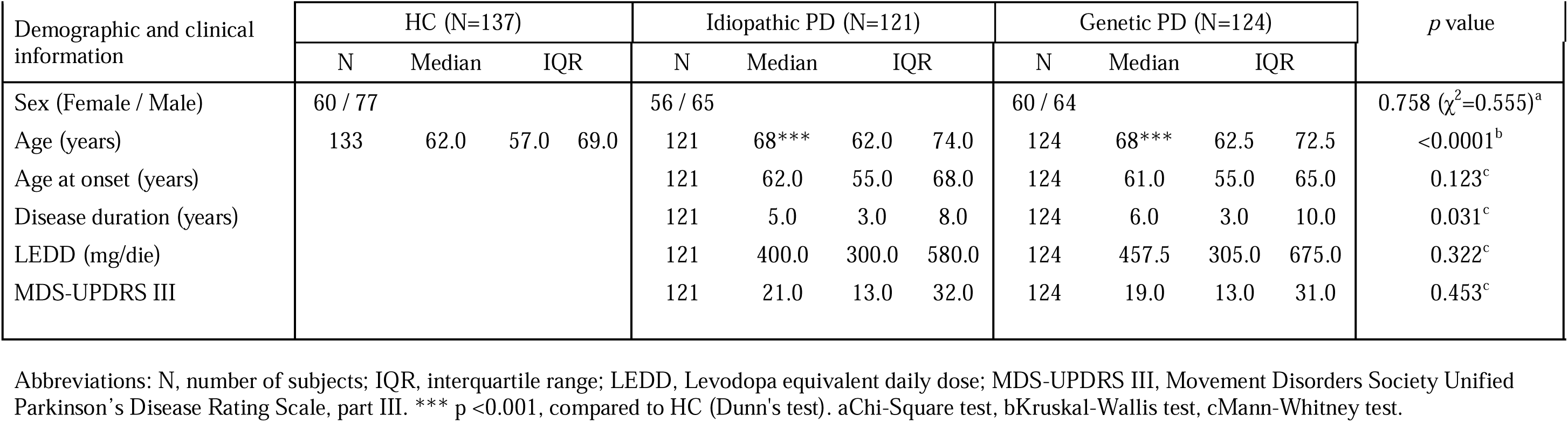
Demographic and clinical characteristics of healthy controls, idiopathic PD and genetic PD enrolled in the serum collection for NMR analysis.

| Demographic and clinical information | HC (N=137) |  |  |  | Idiopathic PD (N=121) |  |  |  | Genetic PD (N=124) |  |  |  | *p* value |
| --- | --- | --- | --- | --- | --- | --- | --- | --- | --- | --- | --- | --- | --- |
|  | N | Median | IQR |  | N | Median | IQR |  | N | Median | IQR |  |  |
| Sex (Female / Male) | 60 / 77 | | | | 56 / 65 | | | | 60 / 64 | | | | 0.758 ($\chi^2$=0.555)<sup>a</sup> |
| Age (years) | 133 | 62.0 | 57.0 | 69.0 | 121 | 68*** | 62.0 | 74.0 | 124 | 68*** | 62.5 | 72.5 | <0.0001<sup>b</sup> |
| Age at onset (years) |  |  |  |  | 121 | 62.0 | 55.0 | 68.0 | 124 | 61.0 | 55.0 | 65.0 | 0.123<sup>c</sup> |
| Disease duration (years) |  |  |  |  | 121 | 5.0 | 3.0 | 8.0 | 124 | 6.0 | 3.0 | 10.0 | 0.031<sup>c</sup> |
| LEDD (mg/die) |  |  |  |  | 121 | 400.0 | 300.0 | 580.0 | 124 | 457.5 | 305.0 | 675.0 | 0.322<sup>c</sup> |
| MDS-UPDRS III |  |  |  |  | 121 | 21.0 | 13.0 | 32.0 | 124 | 19.0 | 13.0 | 31.0 | 0.453<sup>c</sup> |

Abbreviations: N, number of subjects; IQR, interquartile range; LEDD, Levodopa equivalent daily dose; MDS-UPDRS III, Movement Disorders Society Unified Parkinson's Disease Rating Scale, part III. \*\*\* $p < 0.001$ , compared to HC (Dunn's test). <sup>a</sup>Chi-Square test, <sup>b</sup>Kruskal-Wallis test, <sup>c</sup>Mann-Whitney test.

Sex-stratified analyses showed no significant clinical differences between male iPD and gPD patients (**Table S1**). In contrast, female gPD patients showed an earlier age at onset (median, 61.5 vs 64.0 years) and a longer duration of motor symptoms (median, 7.0 vs 5.0 years) than female iPD patients, while age, LEDD, and MDS-UPDRS III scores were comparable between PD subtypes (**Table S2)**.

Among the 124 gPD patients, 30 carried a pathogenic variant in *GBA1* (13 males; 17 females), 17 in *LRRK2* (6 males; 11 females), 40 in *PARK2/PINK1/PARK7* (22 males; 18 females), and 33 in *TMEM175* (21 males, 12 females). Four additional patients carried two PD-associated variants each, involving *GBA1/TMEM175*, *GBA1/PARK2*, GBA1/PINK1 and *TMEM175/PARK2*, respectively. The complete list of variants is reported in **Table S3**.

### NMR metabolomics identifies distinct serum profiles in the whole Parkinson’s disease cohort

We performed comprehensive serum metabolomic profiling by ¹H NMR across the entire cohort, comprising 245 PD patients and 137 sex-matched HCs. Quantitative analysis of 1D ¹H Carr- Purcell-Meiboom-Gill (CPMG) NMR spectra enabled the identification of 43 circulating metabolites (**Fig. S1**).

To determine whether PD was associated with disease-specific serum metabolic signatures, we performed Partial Least Squares Discriminant Analysis (PLS-DA), which showed a clear separation between PD patients and HCs (**Fig. 1A**), with robust predictive performance (Q2 = 0.65, R2 = 0.67 for PC1). The discriminatory capacity of the model was further supported by complementary analytical approaches, including unsupervised Principal Component Analysis (PCA), Receiver Operating Characteristic Area Under the Curve (ROC-AUC), Support Vector Machine (SVM) and Random Forest (**Fig S2; Table S4**).

**Figure 1.**
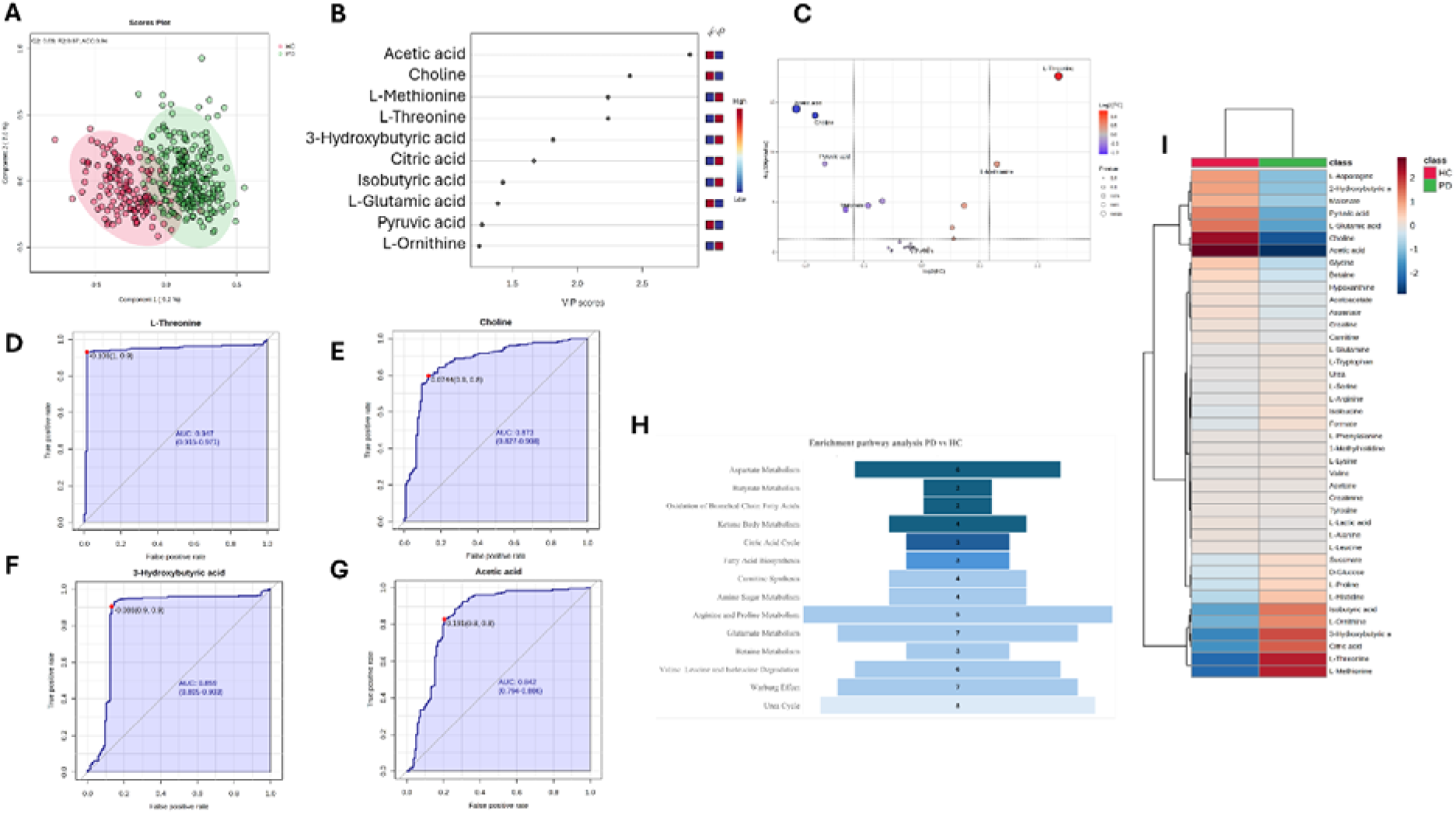
NMR-based serum metabolomics reveals a distinct metabolic fingerprint differentiating Parkinson’s patients from healthy controls. **A**, PLS-DA score plot is shown in Cartesian space, with the x- and y-axes representing the percentage of variance explained by the first and second components (PC1 = 9.2%, PC2 = 7.6). The supervised models were generated using serum metabolite concentrations from 245 Parkinson’s disease (PD) patients versus 137 healthy controls (HCs). Model performance was assessed through 10-fold cross-validation, and R², Q², and accuracy values are reported within each score plot. **B**, Variable Importance in Projection (VIP) scores identifying metabolites contributing to group separation; only metabolites with VIP > 1 were considered relevant. **C**, Robust volcano plots highlighting upregulated (red) and downregulated (blue) metabolites in PD patients relative to HCs, using a fold-change threshold of 1.5 and a p-value < 0.05. **D-G**, Receiver Operating Characteristic (ROC) curves evaluating biomarker performance, where the x-axis represents the false-positive rate and the y-axis the true-positive rate. Each ROC curve includes the empirical ROC curve and the chance diagonal (45° line from (0, 0) to (1, 1)). **H**, Pathway enrichment analyses based on ^1^H-NMR metabolomics data from PD patients compared with HCs. Bars represent the number of detected metabolites (hits) mapped to each pathway. Pathways were considered significant when hits ≥ -2, p < 0.05, Holm-adjusted p and FDR < 0.05. Darker colours indicate lower p-values. Analyses were performed using the Small Molecule Pathway Database (SMPDB) with Homo sapiens selected as the reference organism. **I**, Heatmap of serum metabolite concentrations in PD patients compared with HCs. The colour of each section corresponds to a concentration value of each metabolite calculated from a normalised concentration matrix (red, upregulated; blue, downregulated).

Variable Importance Projection (VIP) analysis identified acetic acid, 3-hydroxybutyric acid, isobutyric acid, citric acid, and pyruvic acid as the principal metabolites differentiating PD patients from HCs (**Fig. 1B**). Alterations in amino acid metabolism also contributed to discrimination between diagnosis groups, with L-methionine, L-threonine, L-glutamic acid, and L-ornithine ranking among the most discriminative metabolites, along with choline (**Fig. 1B**).

Consistent with the multivariate analysis, univariate robust volcano plots showed significant decreases in acetic acid, choline, pyruvic acid, and malonate, together with increases in L-threonine and L-methionine in PD patients compared with HCs (**Fig. 1C; Table S5**).

ROC curve analysis highlighted the strong discriminative power of L threonine (AUC = 0.95), choline (AUC = 0.87), 3-hydroxybutyric acid (AUC = 0.86), and acetic acid (AUC = 0.84) (**Fig. 1D-G**).

To investigate the biochemical processes underlying these metabolic alterations, pathway enrichment analysis was performed on the NMR dataset. This analysis revealed widespread alterations in amino acid homeostasis in PD patients compared with HCs, with aspartate metabolism emerging as the most significantly enriched pathway (**Fig. 1H; Table S6**). Additional changes involved butyrate and ketone body metabolism, branched-chain fatty acid oxidation, fatty acid biosynthesis, the citric acid cycle and the urea cycle (**Fig. 1H; Table S6**). Hierarchical clustering further confirmed the presence of a distinct metabolic signature distinguishing PD patients from HCs (**Fig. 1I**).

Overall, these findings confirm systemic metabolic dysregulation in PD, in which coordinated alterations of individual metabolites converge on interconnected pathways involved in amino acid^42–44^, lipid ^16,45,46^ and energy metabolism ^37,39,42,47,48^, together with urea cycle dysregulation ^49^.

### Sex differences shape serum metabolomic profiles across the Parkinson’s disease cohort

To further investigate whether PD-associated serum metabolomic alterations differed between males and females, we performed sex-stratified analyses comparing PD patients with sex-matched HCs, irrespective of PD subtype. Serum metabolomic profiles from 129 male and 116 female patients with PD were compared with those of 77 male and 60 female HCs.

PLS DA revealed a clear separation between PD and HC serum metabolomes in both sexes (males: PC1 Q² = 0.77, R² = 0.80; females: PC1 Q² = 0.66, R² = 0.61; **Fig. 2A, H**). The robustness of this separation was further supported by complementary unsupervised and supervised approaches (**Fig. S3; Table S4**).

**Figure 2.**
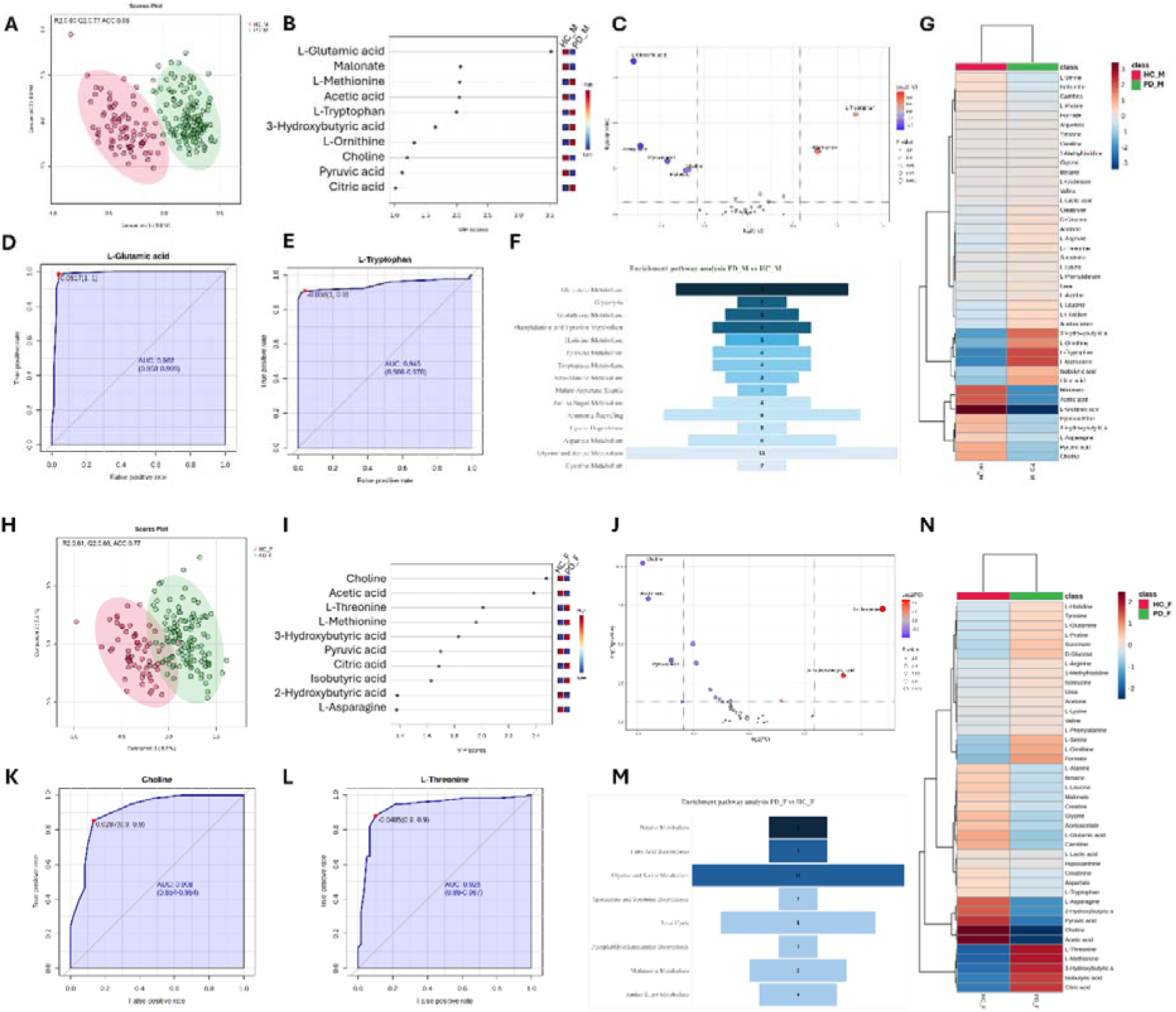
^1^H NMR serum metabolomics shows sex-dependent metabolic profiles in Parkinson’s disease. **A, H,** PLS-DA score plots are shown in Cartesian space, with the x- and y-axes representing the percentage of variance explained by the first and second components (Males: PC1 = 9.8%, PC2 = 8.8%; Females: PC1 = 8.2%, PC2 = 7.5%). The supervised models were generated using serum metabolite concentrations from 129 male Parkinson’s disease (PD) patients versus 77 sex-matched healthy controls (HCs), and from 116 female PD patients versus 60 sex-matched HCs. Model performance was assessed through 10-fold cross-validation, and R², Q², and accuracy values are reported within each score plot. **B, I,** Variable Importance in Projection (VIP) scores identifying metabolites contributing to group separation; only metabolites with VIP > 1 were considered relevant. **C, J,** Robust volcano plots highlighting upregulated (red) and downregulated (blue) metabolites in PD patients relative to sex-matched HCs, using a fold-change threshold of 1.5 and a p-value < 0.05. **D,E,K,L,** Receiver Operating Characteristic (ROC) curves evaluating biomarker performance, where the x-axis represents the false-positive rate and the y-axis the true-positive rate. Each ROC curve includes the empirical ROC curve and the chance diagonal (45° line from (0, 0) to (1, 1)). **F, M,** Pathways enrichment analyses based on ^1^H-NMR metabolomics data from male and female PD patients compared with their respective HCs. Bars represent the number of detected metabolites (hits) mapped to each pathway. Pathways were considered significant when hits ≥ 2, p < 0.05, Holm-adjusted p and FDR < 0.05. Darker colours indicate lower p-values. Analyses were performed using the Small Molecule Pathway Database (SMPDB) with Homo sapiens selected as the reference organism. **G, N,** Heatmap of serum metabolite concentrations in male (**G**) and female (**N**) PD patients compared to sex-matched HCs. The colour of each section corresponds to a concentration value of each metabolite calculated from a normalised concentration matrix (red, upregulated; blue, downregulated).

VIP analysis in males identified several amino acids among the strongest contributors to group separation, with L glutamic acid, L methionine, L tryptophan and L ornithine emerging as the most discriminative biomolecules (**Fig. 2B**). Metabolites related to energy metabolism, including malonate, 3 hydroxybutyric acid, pyruvic acid and citric acid, also contributed substantially to diagnosis groups separation, together with choline (**Fig. 2B**). In contrast, female patients exhibited a distinct metabolic profile, with choline as the strongest contributor, followed by acetic acid, pyruvic acid, citric acid, isobutyric acid, 3-hydroxybutyric acid, and 2-hydroxybutyric acid. Moreover, changes in L-threonine, L-methionine, and L-asparagine also contributed to the separation of serum metabolomes between PD patients and HCs (**Fig. 2I**).

Univariate analyses corroborated these multivariate observations. In males, robust volcano plot analysis revealed significantly reduced circulating levels of L-glutamic acid, acetic acid, pyruvic acid, choline, and malonate, together with increased L-tryptophan and L-methionine, compared with sex-matched HCs (**Fig. 2C; Table S7**). In females, significant reductions in choline, acetic acid, and pyruvic acid were accompanied by elevated 3-hydroxybutyric acid and L-threonine (**Fig. 2J; Table S7**). Hierarchical clustering further supported the presence of sex-specific metabolic signatures distinguishing PD patients from HCs (**Fig. 2G, N**).

ROC analysis identified L-glutamic acid (AUC = 0.98) and L-tryptophan (AUC = 0.94) as the metabolites with the greatest discriminative power in males (**Fig. 2D, E**), whereas choline (AUC = 0.91) and L-threonine (AUC = 0.93) showed the highest discriminative performance in females (**Fig. 2K, L**).

Pathway enrichment analysis indicated that the male PD profile was characterized predominantly by broad alterations in amino acid metabolism, including glutamate, tryptophan, aspartate, and glycine and serine metabolism (**Fig. 2F; Table S8**). Additional alterations involved glutathione metabolism, glycolysis, the malate–aspartate shuttle, and ammonia recycling, indicating coordinated impairments in amino acid, energy, nitrogen and redox homeostasis (**Fig. 2F; Table S8**). In females, the altered metabolites mapped primarily to lipid-, one-carbon- and methyl-donor metabolic processes, including fatty acid and phosphatidylethanolamine biosynthesis, and methionine and betaine metabolism, together with alterations in the urea cycle (**Fig. 2M; Table S8)**.

We next directly compared male and female patients within the whole PD cohort to assess whether sex-specific metabolic signatures were detectable independently of HCs. PLS-DA showed a clear separation between male and female serum metabolomic profiles (R² = 0.56, Q² = 0.51; **Fig. S4A**), which was further supported by PCA, ROC-AUC, SVM, and Random Forest analyses (**Fig. S5; Table S9**). VIP analysis identified L-glutamic acid, L-serine, and L-tryptophan as the principal metabolites discriminating male and female PD patients, together with 2-hydroxybutyric acid, citric acid, L-lactic acid, and carnitine (**Fig. S4B**). Robust volcano plot analysis further showed significantly lower serum L-glutamic acid and L-serine levels in males than in females (**Fig. S4C**), with L-glutamic acid showing the highest discriminative performance (AUC = 0.88; **Fig. S4D**). Finally, pathway enrichment analysis confirmed sex-dependent alterations involving glutamate, tryptophan, glycine and serine metabolism (**Fig. S4E**), while heatmap analysis illustrated the corresponding metabolic patterns (**Fig. S4F**).

To formally quantify the sex dependency of PD-associated metabolic alterations and to account for the potential confounding effect of age, we fitted linear regression models including a group × sex interaction term for the quantified serum metabolites in the full cohort (PD + HC, n = 382). Ten metabolites showed a significant group × sex interaction after FDR correction (p < 0.05; **Table 3**). Because the direction and magnitude of the interaction varied across metabolites, the corresponding coefficients and metabolic trends are summarised in **Table 3**. These findings indicate that the observed sex-dependent metabolic alterations were not attributable to age differences between groups.

**Table 3.** Metabolites showing a significant group × sex interactions across the three regression models.

| Metabolite | PD (all) vs HC<br>(n=245 vs N=133) | iPD vs HC<br>(n=121 vs N=133) | gPD vs HC<br>(n=124 vs N=133) |
| --- | --- | --- | --- |
| L-Tryptophan | $\beta = +0.945$<br>p FDR = 6.70e-27<br>Crossover: F ↓ / M ↑ | $\beta = +1.013$<br>p FDR = 4.32e-32<br>Crossover: F ↓ / M ↑ | $\beta = +0.877$<br>p FDR = 7.58e-10<br>Crossover: F ↓ / M ↑ |
| Formate | $\beta = -0.417$<br>p FDR = 0.0043<br>Crossover: F ↑ / M ↓ | $\beta = -0.428$<br>p FDR = 0.0174<br>Crossover: F ↑ / M ↓ | $\beta = -0.390$<br>p FDR = 0.0233<br>Crossover: F ↑ / M ↓ |
| Tyrosine | $\beta = -0.303$<br>p FDR = 0.0043<br>Crossover: F ↑ / M ↓ | $\beta = -0.263$<br>p FDR = 0.0174<br>Crossover: F ↑ / M ↓ | $\beta = -0.337$<br>p FDR = 0.0112<br>Crossover: F ↑ / M ↓ |
| L-Serine | $\beta = -0.481$<br>p FDR = 0.0116<br>Crossover: F ↑ / M ↓ | $\beta = -0.557$<br>p FDR = 0.0174<br>Crossover: F ↑ / M ↓ | |
| L-Threonine | $\beta = -0.214$<br>p FDR = 0.0262<br>Crossover: F ↑ / M ↓ | | $\beta = -0.325$<br>p FDR = 0.0126<br>Crossover: F ↑ / M ↓ |
| L-Proline | $\beta = -0.143$<br>p FDR = 0.0262<br>Crossover: F ↑ / M ↓ | | |
| L-Glutamic acid | $\beta = -0.747$<br>p FDR = 5.29e-7<br>Concordant ↓ (M>F) | $\beta = -0.743$<br>p FDR = 4.90e-4<br>Concordant ↓ (M>F) | $\beta = -0.750$<br>p FDR = 7.12e-5<br>Concordant ↓ (M>F) |
| Malonate | $\beta = -0.485$<br>p FDR = 0.0043<br>Concordant ↓ (M>F) | $\beta = -0.441$<br>p FDR = 0.0404<br>Concordant ↓ (M>F) | $\beta = -0.526$<br>p FDR = 0.0092<br>Concordant ↓ (M>F) |
| Isoleucine | $\beta = -0.204$<br>p FDR = 0.0189<br>Concordant ↓ (M>F) | | $\beta = -0.232$<br>p FDR = 0.0361<br>Concordant ↓ (M>F) |
| Pyruvic acid | | $\beta = -0.562$<br>p FDR = 0.0413<br>Concordant ↓ (M>F) | |
| Citric acid | $\beta = -0.238$<br>p FDR = 0.0163<br>Concordant ↑ (F>M) | $\beta = -0.279$<br>p FDR = 0.0174<br>Concordant ↑ (F>M) | |

Only metabolites showing a statistically significant group × sex interaction (FDR < 0.05) in at least one of the three models are reported; empty cells indicate that the interaction was not significant in the corresponding model. $\beta$ represents the age-adjusted HC3-robust interaction coefficient (log scale), and p(FDR) the Benjamini-Hochberg adjusted p-value. Trend classification: *Crossover* indicates opposite PD-associated changes between females (F) and males (M); *Concordant* $\uparrow/\downarrow$ (M>F or F>M) indicates that both sexes changed in the same direction, with a greater magnitude in the indicated sex. Rows shaded in grey identify metabolites significant in all three models. Sample sizes are reported in the column headers (comparison group vs. HC reference group). Abbreviations: PD, all Parkinson's disease patients (idiopathic + genetic); iPD, idiopathic PD; hPD, genetic PD; HC, healthy control.

### Sex differences shape serum metabolomic profiles in idiopathic Parkinson’s disease

We next investigated the influence of gender differences on serum metabolomic alterations in iPD, comparing male and female patients (n=65 males, 56 females) with their respective sex-matched HCs (n=77 males, 60 females). PLS-DA revealed a clear separation between iPD patients and HCs in both sexes (males: PC1 Q² = 0.76, R² = 0.80; females: PC1 Q² = 0.73, R² = 0.78; **Fig. 3A, H**). The robustness of this classification was further supported by the complementary univariate and multivariate analytical approaches described above **(Fig. S6**; **Table S4)**.

**Figure 3.**
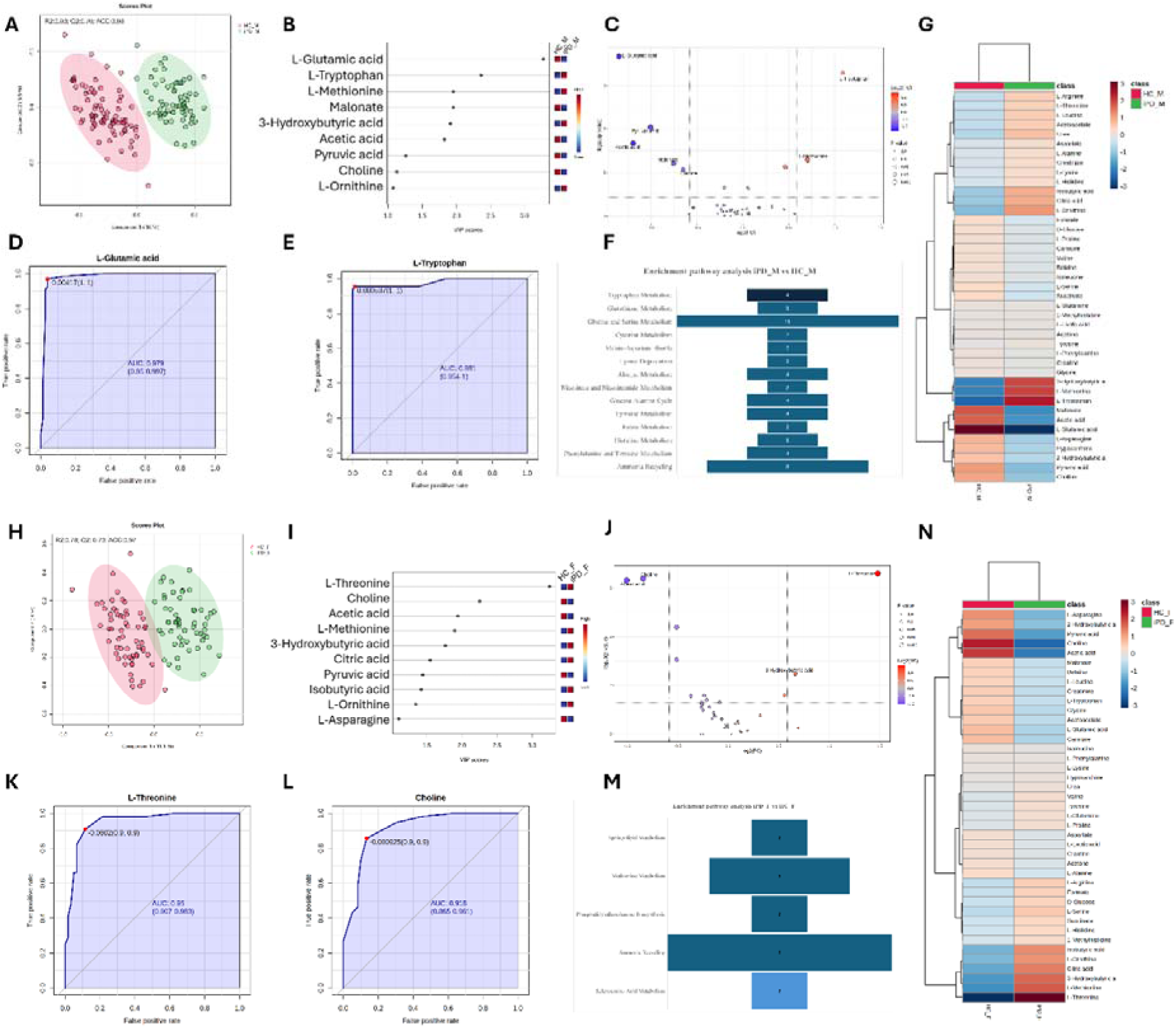
¹H NMR serum metabolomics reveals sex-dependent metabolic profiles in idiopathic Parkinson’s disease compared to HC. **A, H,** PLS-DA score plots are shown in Cartesian space, with the x- and y-axes representing the percentage of variance explained by the first and second components (Males: PC1 = 10%, PC2 = 5.5%; Females: PC1 = 11.1%, PC2 = 4%). The supervised models were generated using serum metabolite concentrations from 65 male idiopathic Parkinson’s disease (iPD) patients versus 77 sex-matched healthy controls (HCs), and from 56 female iPD patients versus 60 sex-matched HCs. Model performance was assessed through 10-fold cross-validation, and R², Q², and accuracy values are reported within each score plot. **B, I,** Variable Importance in Projection (VIP) scores identifying metabolites contributing to group separation; only metabolites with VIP > 1 were considered relevant. **C, J,** Robust volcano plots highlighting upregulated (red) and downregulated (blue) metabolites in iPD patients relative to sex- matched HCs, using a fold-change threshold of 1.5 and a p-value < 0.05. **D,E, K,L,** Receiver Operating Characteristic (ROC) curves evaluating biomarker performance, where the x-axis represents the false-positive rate and the y-axis the true positive rate. Each ROC curve includes the empirical ROC curve and the chance diagonal (45° line from (0, 0) to (1, 1)). **F, M,** Pathway enrichment analyses based on ^1^H-NMR metabolomics data from male and female iPD patients compared with their respective HCs. Bars represent the number of detected metabolites (hits) mapped to each pathway. Pathways were considered significant when Hits ≥ 2, p < 0.05, Holm-adjusted p and FDR < 0.05. Darker colours indicate lower p-values. Analyses were performed using the Small Molecule Pathway Database (SMPDB) with Homo sapiens selected as the reference organism. **G, N** Heatmap of serum metabolite concentrations in male (**G**) and female (**N**) iPD patients compared to sex-matched HC. The colour of each section corresponds to a concentration value of each metabolite calculated from a normalised concentration matrix (red, upregulated; blue, downregulated).

In male iPD patients, VIP analysis identified L-glutamic acid as the major contributor to diagnosis groups discrimination (**Fig. 3B**). Additional amino acids associated with the male metabolic signature included L-tryptophan, L-methionine and L-ornithine (**Fig. 3B**). Biomolecules associated with energy metabolism, including malonate, 3-hydroxybutyric acid, acetic acid, pyruvic acid and choline, also contributed substantially to the separation between iPD patients and HCs (**Fig. 3B**). Conversely, the female iPD metabolomic profile was characterized by prominent contributions from amino acids such as L-threonine, L-methionine, L-ornithine, and L-asparagine, together with 3-hydroxybutyric acid, isobutyric acid, acetic acid, citric acid, and pyruvic acid (**Fig. 3I**). Choline again emerged as one of the principal metabolites discriminating female patients from sex-matched HCs (**Fig. 3I**).

Robust volcano plot analysis revealed significantly reduced serum concentrations of L-glutamic acid, pyruvic acid, acetic acid, malonate and choline, together with increased levels of L-tryptophan and L-methionine, in male iPD patients compared with HCs (**Fig. 3C; Table S10**). Female iPD patients similarly exhibited lower serum levels of choline and acetic acid, accompanied by increased 3-hydroxybutyric acid and L-threonine relative to HCs (**Fig. 3J; Table S10**). Heatmap analysis overall confirmed sex-dependent metabolic differences between iPD patients and HCs (**Fig. 3G, N**).

ROC analysis identified L-glutamic acid (AUC = 0.99) and L-tryptophan (AUC = 0.98) as the strongest distinguishing metabolites in males **(Fig. 3D, E)**, whereas L-threonine (AUC = 0.95) and choline (AUC = 0.91) exhibited the highest discriminative performance in females **(Fig. 3K, L)**.

Based on the altered metabolites identified in male iPD patients, pathway enrichment analysis revealed broad perturbations in amino acid metabolism (**Fig. 3F**), together with alterations in glutathione metabolism, the glucose–alanine cycle, the malate–aspartate shuttle, and ammonia recycling (**Fig. 3F, Table S11**). In contrast, female iPD patients exhibited dysregulation of methionine metabolism, sphingolipid metabolism, and phosphatidylethanolamine biosynthesis (**Fig. 3M; Table S11**). Notably, ammonia recycling emerged as a shared dysregulated pathway in both sexes, indicating a common disturbance of nitrogen homeostasis in iPD.

We next directly compared male and female patients within the iPD cohort to further define sex- dependent metabolic differences. Multivariate analysis showed a clear separation between male and female serum metabolomic profiles (R² = 0.52, Q² = 0.68; **Fig. S7A**), which was further supported by PCA, ROC-AUC, SVM and Random Forest analyses (**Fig. S8; Table S9**). VIP analysis identified L-glutamic acid as the principal discriminator between male and female patients (**Fig S7B**). Consistent with this observation, robust volcano plot analysis showed a significant reduction in serum L-glutamic acid levels in males compared with females (**Fig. S8C**), and ROC analysis confirmed its high discriminatory performance (AUC = 0.88; **Fig. S7D**). Pathway enrichment analysis further identified glutamate metabolism and glutathione biosynthesis among the most dysregulated pathways distinguishing male and female iPD patients (**Fig. S7E**). Heatmap analysis likewise highlighted distinct sex-dependent metabolic patterns (**Fig. S7F**).

Regression modeling conducted on the combined HC+iPD cohort (n=258) identified multiple metabolites showing significant group × sex interaction after FDR correction (**Table 3**). The direction and magnitude of these interactions varied across metabolites, further supporting the presence of a marked sex-dependent metabolic signature in iPD after adjustment for age.

### Sex differences shape serum metabolomic profiles in genetic Parkinson’s disease

We next investigated whether sex modulates serum metabolomic profiles in gPD patients carrying at least one PD-associated variant in *LRRK2*, *TMEM175*, *PARK2*, *PINK1*, *PARK7*, and *GBA1*, compared with sex-matched HCs. PLS-DA showed a clear separation between gPD patients and sex-matched HCs in both sexes (males: n=64 gPD, 77 HC; PC1, Q2 = 0.77, R2 = 0.81; females: n=60 gPD, 60 HC; PC1, Q2 = 0.74, R2 = 0.80; **Fig. 4A, H**). The robustness of the classification was supported by complementary unsupervised and supervised analytical approaches (**Fig. S9; Table S4).**

**Figure 4.**
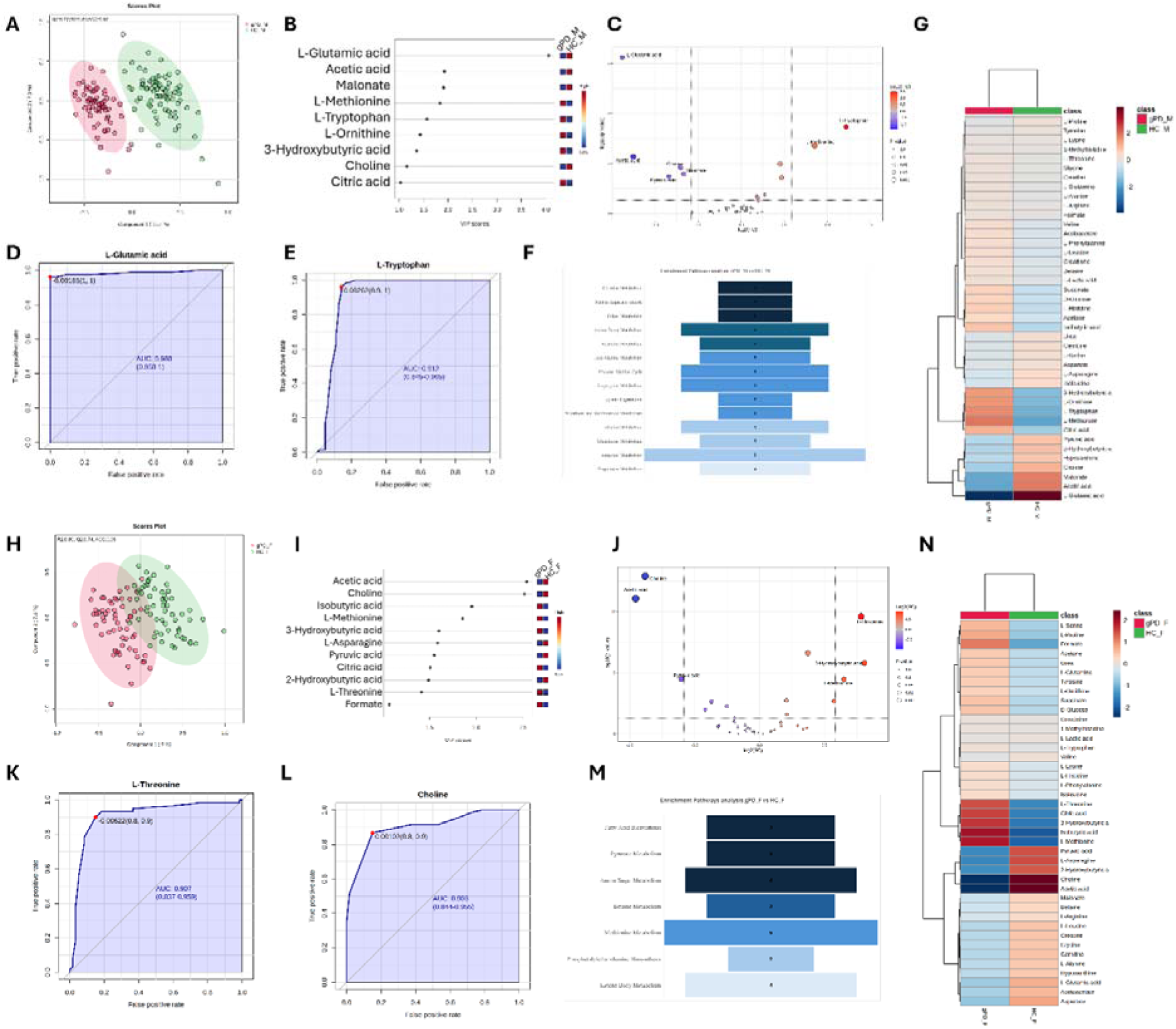
¹H NMR serum metabolomics reveals sex-dependent metabolic profiles in genetic Parkinson’s disease compared to HC. PLS-DA score plots (**Panels A, H)** are shown in Cartesian space, with the x- and y-axes representing the percentage of variance explained by the first and second components (Males: PC1 = 11.7%, PC2 = 7.3%; Females: PC1 = 9 %, PC2 = 7.6%). The supervised models were generated using serum metabolite concentrations from 64 male genetic Parkinson’disease (gPD) patients versus 77 sex-matched healthy controls (HCs), and from 60 female gPD patients versus 60 sex-matched HCs. Model performance was assessed through 10-fold cross-validation, and R², Q², and accuracy values are reported within each score plot. **B, I,** Variable Importance in Projection (VIP) scores identifying metabolites contributing to group separation; only metabolites with VIP > 1 were considered relevant. **C, J,** Robust volcano plots highlighting upregulated (red) and downregulated (blue) metabolites in gPD patients relative to HCs, using a fold-change threshold of 1.5 and a p-value < 0.05. **D,E,K,L,** Receiver Operating Characteristic (ROC) curves evaluating biomarker performance, where the x-axis represents the false-positive rate and the y-axis the true-positive rate. Each ROC curve includes the empirical ROC curve and the chance diagonal (45° line from (0, 0) to (1, 1)). **F, M,** Pathway enrichment analyses based on ^1^H-NMR metabolomics data from male and female gPD patients compared with their respective HCs. Bars represent the number of detected metabolites (hits) mapped to each pathway. Pathways were considered significant when hits ≥ 2, p < 0.05, Holm-adjusted p and FDR < 0.05. Darker colours indicate lower p-values. Analyses were performed using the Small Molecule Pathway Database (SMPDB) with Homo sapiens selected as the reference organism. **G, N,** Heatmap of serum metabolite concentrations in male (**G**) and female (**N**) gPD patients compared to sex-matched HCs. The colour of each section corresponds to a concentration value of each metabolite calculated from a normalised concentration matrix (red, upregulated; blue, downregulated).

In male gPD patients, VIP analysis identified L-glutamic acid as the main discriminative metabolite relative to sex-matched HCs, followed by L-methionine, L-tryptophan, and L-ornithine (**Fig. 4B**). Additional discriminative metabolites included acetic acid, malonate, 3-hydroxybutyric acid, choline and citric acid (**Fig. 4B**). In females, metabolic separation from HCs was mainly driven by acetic acid, isobutyric acid, 3-hydroxybutyric acid, pyruvic acid, citric acid, 2-hydroxybutyric acid and formate, together with choline, L-methionine, L-asparagine, and L-threonine (**Fig. 4I**).

Robust volcano plot analysis confirmed significant downregulations of L-glutamic acid, acetic acid, choline, malonate, and pyruvic acid, along with increased levels of L-tryptophan and L-methionine in male gPD patients compared with sex-matched HCs (**Fig. 4C; Table S12)**. Conversely, female gPD patients exhibited reduced choline, acetic acid and pyruvic acid levels, together with increased L-threonine, 3-hydroxybutyric acid, and L-methionine compared with sex-matched HCs (**Fig. 4J; Table S12**). Hierarchical heatmap analysis further supported sex-specific metabolic signatures distinguishing gPD patients from sex-matched HCs (**Fig. 4G, N**).

ROC analysis identified L-glutamic acid (AUC: 0.98) and L-tryptophan (AUC: 0.91) as the metabolites with the strongest discriminative performance in males (**Fig 4D, E**), whereas L-threonine and choline showed the highest discriminative performance in females (AUC: 0.90 for both; **Fig. 4K, L**).

Based on the altered metabolites identified in male gPD patients, pathway enrichment analysis revealed broad perturbations in amino acid metabolism, including tryptophan, alanine, lysine, and aspartate. Additional alterations involved glutathione metabolism, the glucose-alanine cycle, and the malate-aspartate shuttle, indicating changes in redox and energy homeostasis (**Fig. 4F; Table S13)**. This pathway profile showed substantial overlap with that identified in male iPD patients. In contrast, pathway enrichment analysis in female gPD patients indicated altered fatty acid biosynthesis, pyruvic acid metabolism and phosphatidylethanolamine biosynthesis, together with dysregulation of betaine and methionine metabolism (**Fig 4M**; **Table S13**), closely resembling the pathway profile observed in female iPD patients.

Regression modelling performed on the combined HC+gPD cohort (n=261) identified multiple metabolites showing significant group × sex interactions after FDR correction **(Table 3)**. The direction and magnitude of these interactions varied across biomolecules, indicating the presence of a notable sex-dependent metabolic signature in gPD after adjustment for age.

### Idiopathic and genetic Parkinson’s disease share overlapping serum metabolomic profiles

We next examined whether iPD and gPD patients exhibit distinct serum metabolomic profiles and whether these differences are influenced by sex. PLS-DA did not discriminate iPD from gPD in either males or females (Q² = −0.03 and −0.22, respectively), indicating the absence of predictive metabolic separation between the two PD subtypes (**Fig. 5**).

**Figure 5.**
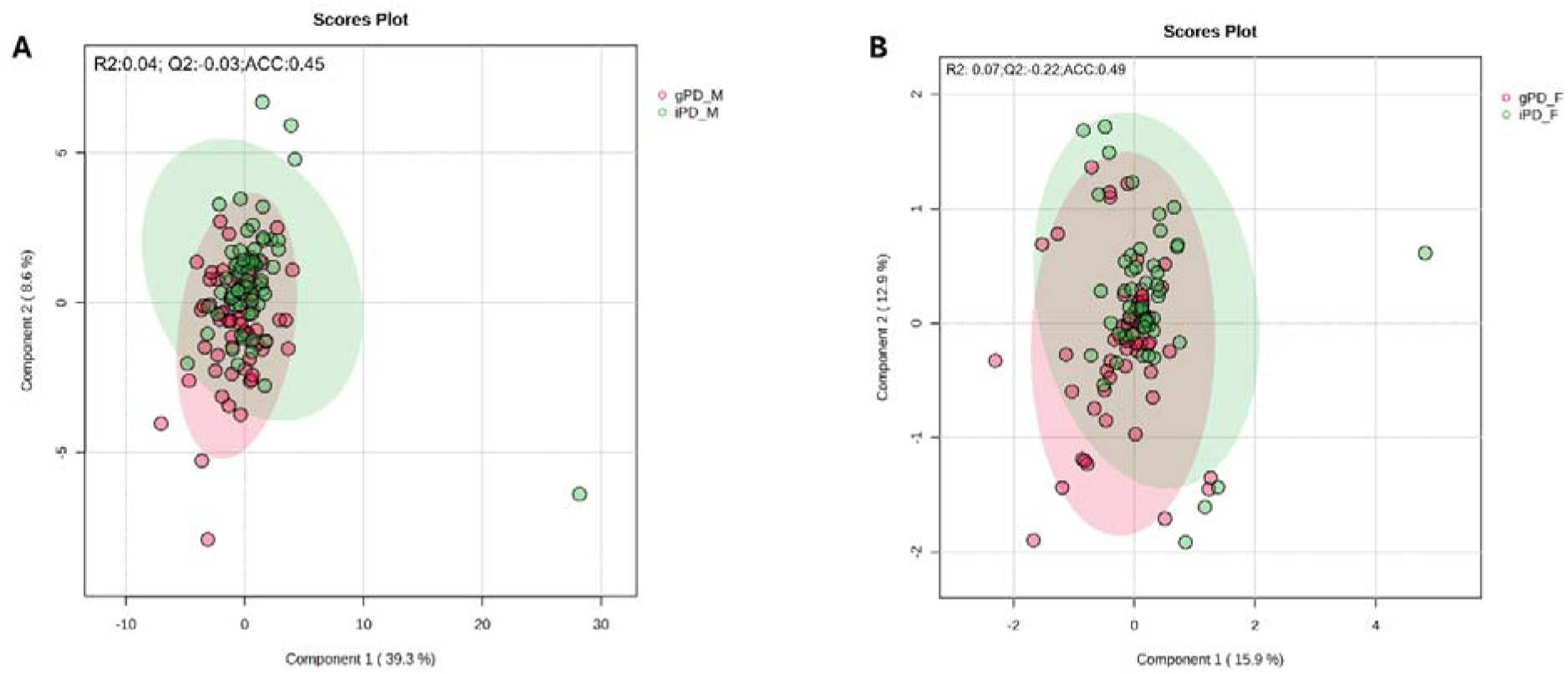
NMR-based metabolomics shows no sex-stratified differences in serum between idiopathic and genetic Parkinson’s subtypes. **A** and **B** are shown in Cartesian space, with the x- and y-axes representing the percentage of variance explained by the first and second components (A: PC1 = 39.3%, PC2 = 8.6%; B: PC1 = 15.9%, PC2 = 12.9%). Supervised models were generated using serum metabolite concentrations from 65 male patients with idiopathic Parkinson’s disease (iPD) and 64 male patients with genetic Parkinson’s disease (gPD), and from 56 female iPD and 60 female gPD patients. Model performance was assessed using 10-fold cross-validation, and R², Q², and accuracy values are reported within each score plot.

To further assess whether subtype-specific differences were detectable after accounting for relevant clinical covariates, linear regression models including a PD subtype × sex interaction term were fitted in the full PD cohort after adjustment for age, disease duration, and LEDD. No metabolite showed a significant PD subtype × sex interaction after FDR correction (all p(FDR) > 0.05), consistent with the multivariate analysis.

Sex-stratified models were then fitted separately in males (n = 129) and females (n = 116) to obtain subtype-specific estimates within each sex. No metabolite differed significantly between iPD and gPD in males after FDR correction. In females, only L-threonine remained significantly associated with PD subtype (β = −0.336, p(FDR) = 0.022), with lower circulating levels in gPD than in iPD.

Overall, these results indicate that iPD and gPD patients share highly overlapping serum metabolomic profiles in both sexes despite their distinct genetic background.

### Integrated metabolomic analysis identifies common and sex-specific metabolic signatures in Parkinson’s disease

To provide an integrated overview of the metabolomic alterations identified across PD subtypes and sexes, we summarised the discriminant metabolites identified by VIP and volcano plot analyses using an UpSet plot (**Fig. 6**).

**Figure 6.**
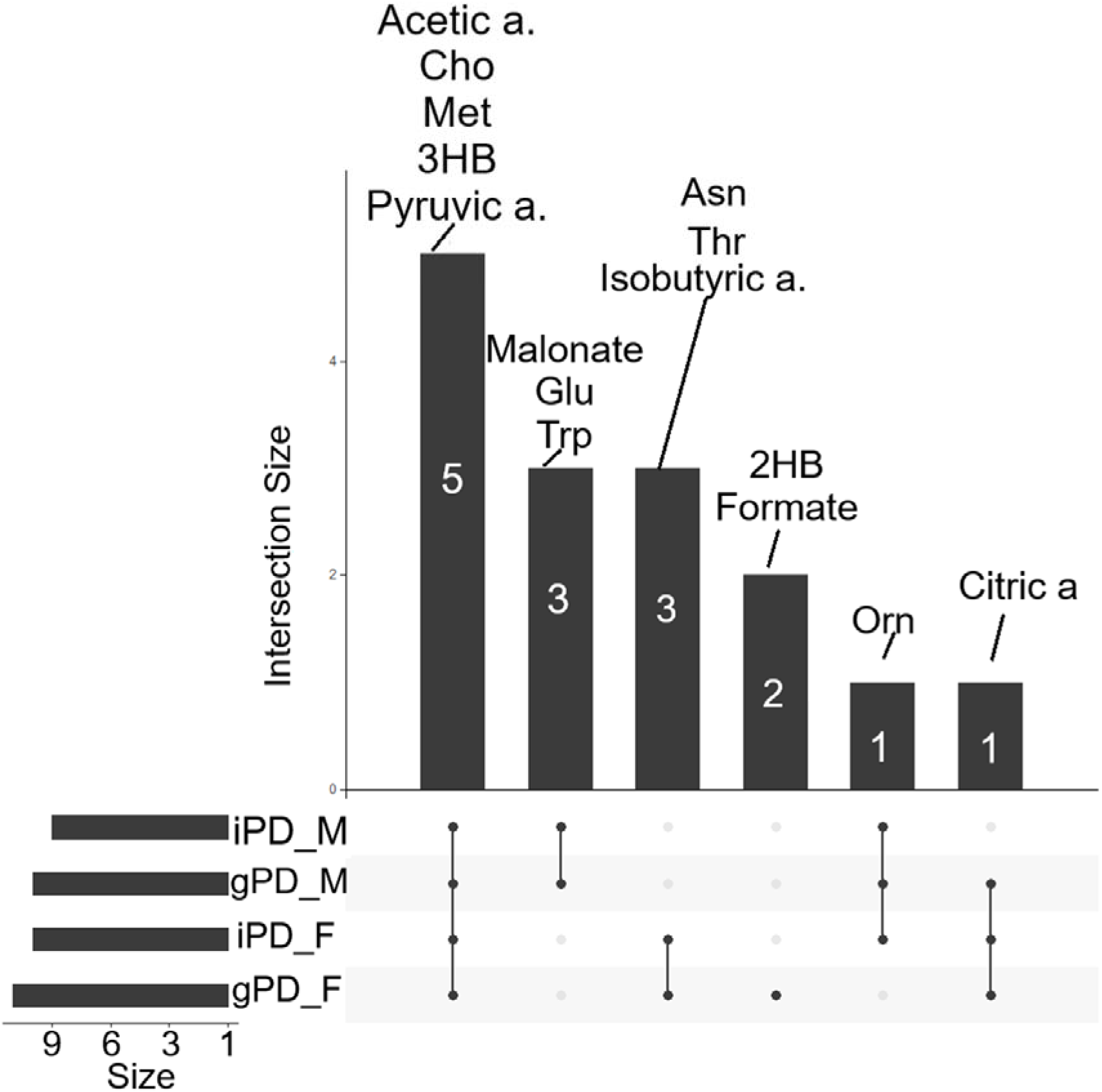
UpSet-based integration of volcano plot and VIP features highlights sex-driven metabolic differences in PD patients. The UpSet plot, created with the Upset R package, displays the overlaps among serum metabolites identified by VIP and robust volcano plot in male and female iPD and gPD groups. The upper bar chart shows the number of metabolites at each intersection; metabolite names are shown above the bars. To improve readability, the names of the discriminating metabolites have been truncated where possible and abbreviated as follows: Acetic a.: acetic acid; Cho: choline; Met: L-methionine, 3HB: 3-Hydroxybutyric acid; Pyruvic a.: pyruvic acid; Asn: asparagine; Thr: L-threonine; Isobutyric a: isobutyric acid; 2HB: 2-hydroxybutyric acid; Orn: L-Ornithine; Citric a: Citric acid.

The UpSet analysis highlighted a core set of discriminant metabolites shared by iPD and gPD patients, irrespective of sex, including reduced circulating levels of choline, acetic acid, and pyruvic acid, together with increased levels of 3-hydroxybutyric acid and methionine (**Fig. 6**). At the same time, the UpSet plot emphasized sex-specific metabolic signatures that were consistently observed across both PD subtypes. In males, PD was characterized by reduced L-glutamic acid and malonate levels, together with increased L-tryptophan, whereas females showed increased L-threonine and isobutyric acid, together with reduced L-asparagine compared with sex-matched HCs (**Fig. 6**; **Figs. 3 and 4**).

Overall, these findings indicate that, although iPD and gPD share a common serum metabolic signature, sex strongly shapes the pattern of discriminant metabolites across PD subtypes.

## Discussion

In the present study, we applied serum ¹H-NMR metabolomics to a large, sex-balanced cohort of 245 clinically characterised PD patients and 137 matched healthy controls, all of whom underwent whole-exome sequencing. This enabled rigorous classification into idiopathic and genetic PD and robust sex-stratified analyses across PD subtypes. Our findings demonstrate that PD is associated with broad alterations in circulating metabolites involved in amino acid, lipid and energy metabolism, consistent with its multisystem nature ^33,38,42,43,45,47–56^. Importantly, complementary multivariate and univariate analyses revealed a marked sex-dependent modulation of these biochemical alterations, resulting in partially distinct metabolic profiles in male and female patients, while idiopathic and genetic PD showed largely overlapping serum metabolomic signatures.

Among the metabolites identified in this study, glutamate emerged as the most consistent hallmark of the male PD metabolic signature. Specifically, both iPD and gPD male patients exhibited a significant reduction in serum glutamate compared with sex-matched HCs. Glutamate is a central metabolic hub involved in protein synthesis, excitatory neurotransmission, immune modulation, nitrogen metabolism, mitochondrial bioenergetics, and redox homeostasis^57–59^. In peripheral tissues, it also contributes to inter-organ nitrogen exchange, hepatic and renal metabolism, pancreatic function, and gastrointestinal signalling^60^. The recurrent reduction in circulating glutamate points to widespread perturbations in amino acid metabolism and energy homeostasis and is in line with previous studies performed in PD cohorts not stratified by sex ^38,42,44,56,61,62^. Furthermore, glutamate is an essential precursor for glutathione biosynthesis, a key component of intracellular redox homeostasis that is impaired in PD ^63–66^. Taken together, these findings support previous evidence indicating a greater impairment of antioxidant defence mechanisms in men with PD ^25,38,42,67–70^. The present NMR findings are consistent with our previous HPLC analyses performed in the same cohort, which likewise identified reduced serum glutamate levels selectively in male PD patients compared with male HCs ^52^.

The convergence of the present NMR findings with our previous HPLC results, together with the sex-dependent pattern consistently observed across PD subtypes and confirmed by linear regression analysis, strengthens the evidence that reduced circulating glutamate is a robust feature of the male PD metabolic profile.

Tryptophan also emerged as a recurrent discriminative metabolite, showing increased circulating levels in male patients with both iPD and gPD, compared with sex-matched HCs. As an essential amino acid, tryptophan is involved in protein synthesis, neurotransmitter synthesis, and immune regulation ^71–75^. Altered tryptophan metabolism has been implicated in PD through the generation of neuroactive metabolites, particularly along the kynurenine pathway, which may contribute to neurodegeneration ^76–78^. Increasing evidence also links gut microbiota dysbiosis to altered tryptophan metabolism in PD and other neurological disorders ^74,79–81^. Although most previous studies reported reduced circulating tryptophan levels in PD^71,77,82^, a recent sex-stratified study identified increased tryptophan levels in male patients and reduced levels in female patients compared with their respective controls^78^. This observation is consistent with our finding of increased circulating tryptophan in male iPD and gPD patients and suggests that tryptophan-related metabolic alterations may differ according to sex in PD.

Importantly, NMR metabolomic analysis revealed reduced serum levels of pyruvate, acetate, malonate, and choline, together with increased methionine levels, in male patients with both iPD and gPD compared with sex-matched HCs. Collectively, these recurrent metabolic alterations, together with the marked reduction in glutamate, point to coordinated disturbances in energy homeostasis, amino acid metabolism, and redox-related processes in male PD patients ^42,53,83–87^.

In contrast, female patients with both iPD and gPD exhibited a distinct metabolic signature characterized by increased serum threonine, reduced choline, and alterations in lipid- and energy- related metabolites compared with sex-matched HCs. Increased circulating threonine has previously been reported in PD, although most studies did not systematically assess sex-specific metabolic effects ^42,56,88^.

Hence, our current NMR findings refine these previous observations by showing that increased serum threonine is a recurrent feature of female patients with both iPD and gPD, suggesting that sex contributes substantially to threonine-related metabolic alterations in this neurological disorder.

Reduced circulating choline was observed in both male and female PD subgroups, indicating that it represents a shared metabolic alteration. However, choline emerged consistently in the female VIP, volcano and ROC analyses, suggesting a greater relative contribution to the female PD metabolic profile. Choline lies at the interface between cholinergic neurotransmission, phospholipid metabolism, membrane integrity, and one-carbon metabolism, acting as a precursor of acetylcholine and phosphatidylcholine as well as a source of betaine for methyl-donor pathways ^89,90^. Previous metabolomic and lipidomic studies have implicated choline-containing phospholipids, including phosphatidylcholine and glycerophosphocholine species, in PD ^91,92^. Notably, a recent sex-stratified metabolomic analysis identified prominent glycerophosphocholine alterations specifically in female PD patients ^93^, consistent with the greater discriminative contribution of choline observed in women in the present study.

Female patients with both iPD and gPD also exhibited significant alterations in serum acetate, isobutyrate, and 3-hydroxybutyrate compared with sex-matched HCs. Acetate is a major gut microbial short-chain fatty acid, whereas isobutyrate is a branched-chain fatty acid associated with microbial protein fermentation ^94–99^. Given the gut microbiota alterations reported in PD^100,101^, reduced circulating acetate may reflect disrupted host–microbiome metabolic interactions, although the origin of this alteration remains to be clarified. In contrast, 3-hydroxybutyrate (β-hydroxybutyrate), the major circulating ketone body generated through fatty acid oxidation, serves as an alternative mitochondrial energy substrate ^102–104^, suggesting adaptations in systemic energy homeostasis.

Based on these metabolite alterations, pathway enrichment analysis further indicated perturbations in phosphatidylethanolamine biosynthesis across female PD subgroups, whereas fatty acid biosynthesis was more prominent in female gPD patients, and sphingolipid metabolism emerged specifically in female iPD patients. Collectively, these findings indicate that the female PD metabolic profile involves coordinated changes linking lipid remodelling, one-carbon metabolism, and gut microbiota-related metabolism.

Despite the recognized genetic and clinical heterogeneity of PD^105,106^, our NMR analyses demonstrated substantial overlap between the serum metabolomic profiles of iPD and gPD, even after sex stratification. Within the metabolite coverage and analytical resolution of the present NMR approach, the absence of distinct clustering suggests that shared disease-related processes predominate over genotype-specific circulating signatures. This interpretation is consistent with our previous observation of largely overlapping metabolic profiles between iPD patients and carriers of rare, non-pathogenic variants in PD-associated genes^51^.

Nevertheless, pathway enrichment analyses suggested some qualitative differences between PD subtypes when each group was compared separately with sex-matched HCs. In particular, amino acid-related perturbations appeared broader in male iPD than in male gPD patients when each group was compared with sex-matched HCs. In particular, male iPD patients showed broader perturbations in glycine/serine, phenylalanine, and tyrosine metabolism. Notably, this observation is consistent with our previous HPLC analyses in the same cohort, which identified lower circulating levels of several neuroactive amino acids in male iPD patients than in male carriers of PD-related genetic variants^52^. Together, these findings suggest a possible interplay among neuroactive amino acid metabolism, sex, and PD subtype that warrants further investigation.

Although the lack of a drug-naïve condition prevents us from fully excluding a potential influence of dopaminergic treatment on the serum metabolome features, the comparable LEDD and disease duration between male and female patients within each PD subtype suggest that the identified sex- related differences are not primarily driven by treatment exposure or disease chronicity. This interpretation is further supported by previous metabolomic studies showing that systemic alterations, including amino acid dysregulation, are already detectable in *de novo* or drug-naïve PD patients ^38,107,108^ . In this respect, serum NMR profiling revealed sex-related metabolic signatures in *de novo* drug-naïve PD patients, with a more pronounced disease-associated fingerprint in males^38^. Consistently, metabolomic analyses comparing treated and drug-naïve PD patients, as well as large- scale plasma profiling studies including untreated *de novo* cases, indicate that at least part of the circulating metabolic dysregulation observed in PD may occur independently of pharmacological treatment and long-term disease progression ^38,107,108^. Moreover, targeted analyses of circulating glutamatergic system-related amino acids recently showed that male patients with early-stage, L- DOPA–naïve PD already display altered amino acid levels, further supporting the notion that amino-acid dysmetabolism may arise early in the disease course.

To further account for potential clinical confounders, regression models simultaneously adjusted for age, disease duration and LEDD identified no significant PD subtype × sex interaction after FDR correction. Likewise, sex-stratified analyses identified only one significant subtype-specific difference -lower threonine in female gPD than in female iPD patients- with no significant differences in males. These results reinforce the overall metabolic convergence between PD subtypes and indicate that age, disease duration and LEDD did not materially account for the observed sex-dependent patterns.

Regression and UpSet analyses provided complementary information on the shared and sex-dependent components of the PD metabolomic profile. Notably, glutamate showed the largest and most significant group × sex interaction across the three disease-control comparisons (overall PD versus HC, iPD versus HC, gPD versus HC) after adjustment for age, further supporting its role as the principal metabolite underlying the male metabolic signature. Malonate, L-tryptophan, tyrosine and formate also exhibited significant group × sex interactions across all the comparisons, indicating that sex-dependent metabolic differences extend beyond glutamate to involve multiple circulating metabolites. The UpSet analysis identified five recurrent discriminant metabolites shared across male and female PD patients: acetate, choline, methionine, 3-hydroxybutyrate and pyruvate. In addition, glutamate, malonate and tryptophan formed a recurrent male-associated set, whereas asparagine, threonine and isobutyrate were shared in females as discriminative metabolites. Together, these findings indicate that PD is characterized by a common metabolic component accompanied by distinct sex-dependent metabolic patterns, supporting sex as a major biological determinant of systemic metabolic remodelling in PD.

Although our findings demonstrate robust sex-associated alterations in circulating metabolism, it remains uncertain to what extent these changes directly reflect metabolic dysfunction within basal ganglia circuits. Rather, serum metabolite levels may be shaped by the interplay among central neurodegeneration, peripheral organ metabolism, diet and gut-associated processes. This issue is particularly relevant for neuroactive D- and L-amino acids, which participate in neurotransmission or neuromodulation but whose circulating levels are also associated with liver and kidney function, lipid and glucose homeostasis ^109–115^. The observed serum profiles should therefore be interpreted as systemic metabolic readouts rather than direct proxies of brain metabolism, potentially integrating contributions from peripheral organ dysfunction and gut dysbiosis in PD.

A major strength of our study is its integrated analytical design, combining extensive serum NMR metabolomic profiling with rigorous clinical, genetic and sex stratification of a large PD cohort. An additional strength is that all statistical comparisons were performed using sex-matched groups of genetically characterised iPD and gPD patients and HCs. Importantly, all participants underwent Whole Exome Sequencing (WES), enabling accurate classification into iPD and gPD while excluding PD-associated pathogenic or likely pathogenic variants from the iPD and HC groups. This rigorous molecular characterisation strengthens the interpretation of the observed metabolic convergence between iPD and gPD.

Different limitations should also be acknowledged. First, the cross-sectional design precludes causal inference regarding the relationships among metabolic alterations, sex differences and PD subtypes. Second, grouping carriers of different PD-associated variants may have masked gene-specific metabolic signatures. Third, although NMR provides highly reproducible and quantitative measurements, its lower sensitivity and more limited metabolite coverage than mass spectrometry may have constrained our ability to detect subtle genotype-specific alterations. Finally, the exclusive analysis of serum may not fully capture metabolic changes restricted to specific tissues or biological matrices. Validation in larger, independent cohorts that integrate complementary metabolomic platforms and multiple biological matrices will therefore be essential to characterize the systemic metabolic landscape of PD further.

Overall, our findings identify biological sex as a major determinant of circulating metabolic alterations in PD, whereas idiopathic and genetic forms largely converge towards a common systemic metabolic phenotype. These observations may help explain part of the heterogeneity reported across previous metabolomic studies ^37^ and underscore the importance of incorporating sex stratification into the design, analysis and interpretation of future metabolomic and biomarker studies in PD.

## Methods

### Study cohort

This is a case-control observational study. The study included 245 independent and unrelated patients with PD who were enrolled for both NMR metabolomic profiling and genetic analyses. These patients were genetically classified into 121 idiopathic and 124 genetic PD patients carrying at least one pathogenic mutation in the most frequently mutated genes *LRRK2*, *GBA1*, *TMEM175* and *PARK2/PINK1/PARK7* (see next paragraph “Genetic mutation analysis of the study cohort” for details about the patient selection).

These patients were part of the PD biobank of IRCCS Neuromed/IGB-CNR. The mean age at diagnosis was 66.5 years (Standard deviation (SD) 8). All subjects were of European ancestry and were evaluated by qualified neurologists at the Parkinson Centre of the IRCCS INM Neuromed from June 2015 to December 2017 and from June 2021 to December 2023, using a thorough protocol comprising a neurological examination and evaluation of non-motor domains. Information about family history, demographic characteristics, anamnesis, and pharmacological therapy was also collected (the treatment of the PD groups consisted, for the most part, of a combination of levodopa and dopamine agonist) ^19^. Clinical criteria for diagnosis required the presence of at least two cardinal motor signs: asymmetric resting tremor, bradykinesia and rigidity, as well as a good response to levodopa and absence of other atypical features and causes of parkinsonism. Exclusion Criteria for enrolment were: *i*) pre-existing psychiatric conditions; *ii*) presence of neurodegenerative neurological diseases such as multiple sclerosis, amyotrophic lateral sclerosis, Alzheimer’s, neuromuscular pathologies, epilepsy; *iii*) diagnosis of dementia; *iv*) depression; *v*) prolonged intake of anxiolytics, antidepressants, antipsychotics, hypnotic drugs, cognitive stimulants.

The Movement Disorder Society revised version of the Unified Parkinson’s Disease Rating Scale Part III (33 items, maximum score 132; hereafter called MDS-UPDRS III) ^116^was used to assess clinical motor symptoms. These included language, facial expressions, tremor, rigidity, agility in movements, stability, gait and bradykinesia. Patients were analyzed during the ON period.

The study also included 137 HC subjects matched for sex with PD cohort. Only individuals aged ≥40 years were included. All the HCs were negative for mutation/variant in PD genes (see next paragraph “Genetic mutation analysis of the study cohort” for the criteria used for patient selection). To minimize potential sources of bias, standardized protocols for clinical assessment, sample collection, and processing were applied. Laboratory analyses were performed on anonymized samples. Sex-matched controls were included to reduce confounding effects.

All procedures involving human participants were approved by the Institutional Review Board of the IRCCS Neuromed Italy. The study protocols N°9/2015, N°19/2020, N°4/2023 have been registered in clinicaltrial.gov with the numbers NCT02403765, NCT04620980, NCT05721911.

Clinical investigations were conducted according to the principles expressed in the Declaration of Helsinki. Written informed consent was obtained from all participants.

The research was carried out following the recommendations set out in the Global Code of Conduct for Research in Resource-Poor Settings.

### Reporting Standards

"Reporting adheres to the STROBE guidelines for case-control studies and MSI guidelines for metabolomics data."

No formal sample size calculation was performed; the sample size was determined based on the availability of well-characterized subjects from the biobank.

### Genetic mutation analysis of the study cohort

To clarify the impact of the genetic background on the metabolic profile, we enrolled 245 PD patients and 137 HCs (**Table 1**). This cohort was previously analyzed in our recent study focused on HPLC analysis of circulating NMDAR-related amino acids and their precursors^52^ and comprises iPD, gPD patients and HCs. Whole-exome sequencing was performed for all participants and was used to classify patients into iPD and gPD, while excluding PD-associated pathogenic or likely pathogenic variants from the iPD and HC groups. The 121 idiopathic PD patients were selected from the entire cohort as those negative for mutations and rare (Minor allele frequency (MAF) < 0.001), potentially harmful variants (Combined Annotation Dependent Depletion (CADD) score ≥15) in a large number of PD candidate genes including those reported in the literature as Mendelian PD genes (*PARK7, DNAJC13, DNAJC6, EIF4G1, FBXO7, LRRK2, PARK2, PINK1, SNCA, VPS35*) and those reported in our recent studies as PD genes/at risk factors (*AIMP2, ANKK1, ANKRD50, CHMP1A, GBA1, GIGYF2, GIPC1, GRK5, HMOX2, HSPA8, HTRA2, IMMT, KIF21B, KIF24, MAN2C1, PACSIN1, RHOT2, SLC25A39, SLC6A3, SNCAIP, SPTBN1, TMEM175, TOMM22, TVP23A, UCHL1, VPS8, ZSCAN21*)^19,117^.

The second group, named “genetic-PD”, includes 124 PD-affected subjects carrying at least 1 pathogenic mutation in one of the most frequently mutated PD genes which include *GBA1, LRRK2, PARK2, PARK7, PINK1*, and *TMEM175*. In particular, we selected 17 patients mutated in *LRRK2*, 30 mutated in *GBA1*, 40 with at least 1 mutation in *PARK2/PINK1/PARK7*, 33 mutated in *TMEM175*, and four patients carrying two mutations in PD genes such as *GBA1/TMEM175*, *GBA1/PARK2*, GBA1/PINK1 and *TMEM175/PARK2*, respectively (see **Table S3** for the detailed list of mutations/variants). All the HCs that we included in the study were negative for mutation/variant in the selected panel of PD genes.

### Collection and storage of serum samples

Blood sampling was performed after a 6-hour fasting. Whole blood was collected by peripheral venipuncture into clot activator tubes and gently mixed.

Sample was stored upright for 30 min at room temperature to allow blood to clot and centrifuged at 2000 ×g for 10 min at room temperature. Serum was aliquoted (0.5 ml) in polypropylene cryotubes and stored at −80 °C before usage. Unique anonymized codes have been assigned to the samples for processing and subsequent analysis, maintaining the confidentiality of personal data.

### NMR sample preparation

Serum samples were prepared in accordance with the NMR metabolomics guidelines to maintain sample quality ^118^. Blood samples were allowed to clot and were subsequently centrifuged to separate the serum from the cellular fraction. Serum aliquots were subsequently stored at −80 °C until NMR analysis. To prepare NMR samples, 100 μL of phosphate buffer (0.075 M Na_2_HPO_4_·7H_2_O, 4% NaN_3_, and water) was mixed with 100 μL of blood serum and then placed in a 3-mm NMR tube. To calibrate and measure NMR signals, we used trimethylsilyl propionic acid and sodium salt (0.1% TSP in D_2_O) as an internal reference. NMR investigations were conducted using a Bruker DRX600 MHz spectrometer (Bruker, Karlsruhe, Germany) equipped with a 5 mm triple-resonance z-gradient TXI Probe ^119,120^. For spectrometer control and data analysis, we utilised TOPSPIN, version 3.2 (Bruker Biospin, Fällanden, Switzerland).

### NMR sample acquisition

The acquisition of the Carr-Purcell-Meiboom-Gill (CPMG) spectrum was essential because serum contains macromolecules, such as proteins, that may interfere with signals related to other metabolites ^118,121^ CPMG experiments were acquired using a spectral width of 7 kHz and 32k data points. Water presaturation was applied for 5 seconds during the relaxation delay, and a spin-echo spacing of 0.3 ms was employed. The time-domain data were processed using a weighted Fourier transform with a 0.5 Hz line-broadening factor, followed by manual optimisation of phase, pH referencing, and baseline correction prior to targeted profiling.

Metabolite identification was performed using the Chenomx NMR Suite^122^ , which enables spectral deconvolution by comparing spectra with a curated library of reference metabolite signatures. All resonances in the ^1^H NMR spectra were carefully inspected and manually assigned using the Profiler module, which overlays theoretical spectral models onto experimental data and refines parameters such as chemical shift, peak intensity, and line shape. Quantification and manual assignments obtained with Chenomx^122^ were subsequently cross-validated using the automated platform Bayesil ^123^ ensuring consistency and robustness in metabolite annotation.

## Statistical analysis

The primary outcome was serum metabolomic profiles. The main exposures were PD subtype (iPD, gPD) and sex. Covariates included age, disease duration, and L-DOPA equivalent daily dose (LEDD).

Clinical and demographic characteristics were described using, as summary statistics, median and the interquartile range (IQR) or absolute and relative frequencies. Comparisons between PD patients and HC were evaluated using Mann Whitney U test for continuous variables and Chi-Square test for dichotomous variables.

The concentration matrices derived from NMR peak quantifications were examined using a univariate method that integrated a T-test with fold-change analysis, depicted in a robust volcano plot. A fold-change threshold of 1.5 and a p-value threshold of less than 0.05 were established ^124^.

Prior to statistical analysis, metabolite concentration matrices were normalised using sum normalisation followed by Pareto scaling. To avoid the risk of overfitting inherent in supervised approaches, the initial exploration of the dataset was conducted using unsupervised PCA. The robustness of the PCA separation was assessed using Permutational Multivariate Analysis of Variance (PERMANOVA), which was also used to evaluate models comparing more than two clusters ^125^.

Supervised modelling was subsequently carried out using PLS-DA implemented in MetaboAnalyst 6.0 and validated using AUC and support vector Machine ^126^. Performance was assessed using the R² and Q² coefficients derived from 10-fold internal cross-validation, reflecting the explained and predicted variance, respectively ^127,128^.

Positive values of both metrics were considered indicative of meaningful model structure, and overall classification accuracy was also reported.

Additional classification analyses were performed using Random Forest. Model quality was evaluated through the out-of-bag (OOB) error rate, class-specific error rates, and explicit reporting of correctly and incorrectly classified samples ^129,130^

Metabolites contributing most strongly to group discrimination were ranked according to their Variable Importance in Projection (VIP) scores. VIP values, calculated as weighted sums of squared PLS-DA loadings, were considered relevant when exceeding 1. To visualise group-level metabolic patterns, heatmaps were generated using normalised concentrations, group means, and Euclidean distance as the similarity metric^131^. Biomarker performance was assessed by analysing the univariate ROC curves, from which the AUC values and their 95% confidence intervals were derived. Confidence intervals were estimated using 500 bootstrap resampling cycles, providing a robust evaluation of classifier stability ^126^.

Pathway analysis was conducted using the Enrichment module of MetaboAnalyst 6.0, applying the global test algorithm for metabolite set enrichment analysis (MSEA)^126^. The Small Molecules Pathways Database (SMPDB) for *Homo sapiens* ^132^ was selected due to its integration with the Human Metabolome Database, including serum-specific metabolite entries derived from NMR spectroscopy ^133^. The analysis incorporated a background list comprising all metabolites detected in the spectra. Only pathways meeting the criteria of false discovery rate (FDR) and adjusted p-value < 0.05 and containing at least two matched metabolites were retained. An UpSet plot was generated using the UpSetR package to visualise intersections among significant discriminating metabolites detected by the Volcano plot and VIP^134^.

To formally test whether the association between PD diagnosis and serum metabolite concentrations differed by sex, and to account for potential confounding by age, we complemented the multivariate analyses with a series of linear regression models. All metabolite concentrations were natural log- transformed prior to modelling. Models were estimated by ordinary least squares (OLS); since the Breusch-Pagan test indicated significant heteroscedasticity in the residuals of a proportion of models, heteroscedasticity-consistent robust standard errors (HC3) were used for all inference, yielding valid test statistics without requiring homoscedasticity of the residuals. A model was fitted in the full cohort (PD + HC) separately for each of the 41 NMR-quantified metabolites (excluding methanol and ethanol, which were considered contaminants), taking the form: log(metabolite) ∼ group + sex + age + group:sex, where group = 0 (HC) or 1 (PD), sex = 0 (female) or 1 (male), and age was a continuous covariate. Identical models were fitted in the HC+iPD and HC+gPD subsamples separately. The group × sex interaction coefficient (β) quantifies whether the PD-vs-HC difference in log-metabolite levels is larger in males than in females; a negative β indicates a greater reduction associated with PD in males relative to females.

To investigate whether metabolic differences between iPD and gPD were modulated by sex within the PD cohort, we first fitted a model of the form: log(metabolite) ∼ group-PD + sex + age + DD + LEDD + group-PD:sex in the full PD subsample. Prior to modelling, collinearity among covariates was assessed using the variance inflation factor (VIF); age at onset was excluded due to high collinearity with age and disease duration (VIF > 10), while all retained covariates showed VIF < 1.4. Since no groupPD × sex interaction reached statistical significance after FDR correction, sex-stratified models of the form log(metabolite) ∼ groupPD + age + DD + LEDD were subsequently fitted separately in females and males to obtain sex-specific estimates of the iPD vs gPD difference without imposing the assumption of equal effects across sexes.

For all models, raw p-values for the interaction term (or for the group PD term in stratified models) were corrected for multiple comparisons using the Benjamini-Hochberg false discovery rate (FDR) procedure across all 41 metabolite outcomes. All analyses were performed in Python using the statsmodels library (v0.14).

## Declarations Data availability

Metabolomics data have been deposited to the EMBL-EBI MetaboLights database (https://www.ebi.ac.uk/metabolights/) with the identifier **MTBLS11187**.

## Code Availability

Not applicable

## Acknowledgments

The authors are grateful to all the patients, their caregivers, the Clinical Parkinson’s Disease Center of IRCCS Pozzilli and the PD biobank of IRCCS Neuromed and IGB-CNR for the kind cooperation with this study.

The work of T.E. was supported by Next Generation EU - PNRR M6C2 Investimento 2.1 valorizzazione e potenziamento della ricerca biomedica del SSN grant n. PNRR-MCNT2-2023- 12377375. TE was also supported by Ministry of Health, Ricerca Corrente. The study of TE was partially funded by Ministry of Enterprises and Made in Italy (MIMIT) project Neurotechno n. F/180029/01/X43 and by Progetto MARADONA, finanziato nell’ambito dell’Accordo per la Coesione della Regione Campania, fondo di rotazione ex l. 183-1987, avviso "Malattie Rare".

This study was also partially funded by Italian Ministry of University and Research (PRIN 2022 - COD. 2022XF7YYL_02 to AU and PRIN 2022 – COD. 2022W3RKLJ to TE). The work of A.U., T.N. and T.E. was supported by NEXTGENERATIONEU (NGEU) and funded by the Ministry of University and Research (MUR), National Recovery and Resilience Plan (NRRP), project MNESYS (PE0000006) – A Multiscale integrated approach to the study of the nervous system in health and disease (DN. 1553 11.10.2022).

The funders had no role in study design, data collection and analysis, decision to publish, or preparation of the manuscript.

## Author contributions

CM: Investigation, data analysis, review & editing; FC: data analysis, review & editing; TN: data analysis, review & editing; MV: Data analysis, Writing –review & editing; MS: Writing –review & editing; SP: review & editing; NM: review & editing; MG: review & editing; AMD: data analysis, review & editing; FE: Writing – review & editing; TE; Funding acquisition, Project administration, Supervision, Writing – review & editing. AU: Conceptualization, Funding acquisition, Project administration, Supervision, Writing – review & editing.

