## Supplementary data for "Gender-associated differences in serum metabolite profiles among patients with Parkinson’s Disease"

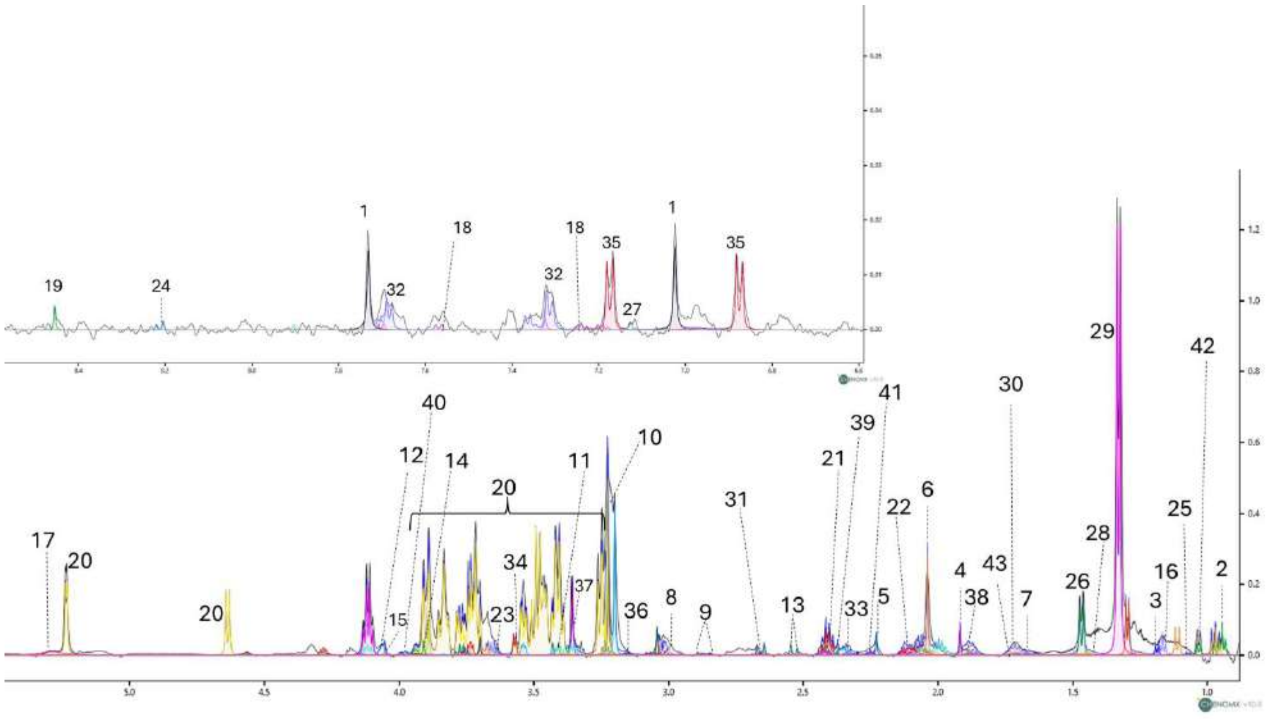


**Figure S1. 1D $^1$H-CPMG spectrum related to PD serum**

1: 1-Methylhistidine; 2:2-Hydroxybutyric acid; 3:3-Hydroxybutyric acid; 4:Acetic acid; 5: Acetoacetic acid; 6: Acetone; 7: Arginine; 8: L-Asparagine; 9: Aspartate; 10:Betaine; 11: Carnitine; 12:Choline; 13: Citric acid; 14: Creatine; 15:Creatinine; 16:Ethanol; 17: Urea; 18: L-Tryptophan; 19: Formate; 20: D-Glucose; 21:L-Glutamic acid; 22: L-Glutamine; 23: Glycine; 24: Hypoxanthine; 25: Isobutyrate; 26: L-Alanine; 27: L-Histidine; 28: Isoleucine; 29: L-Lactic acid; 30: L-Lysine; 31:L-Methionine; 32: L-Phenylalanine; 33: L-Proline; 34: L-Threonine; 35: Tyrosine; 36: Malonate; 37:Methanol; 38: L-Ornithine; 39:Pyruvic acid; 40: L-Serine; 41: Succinate, 42:Valine; 43:L-Leucine.

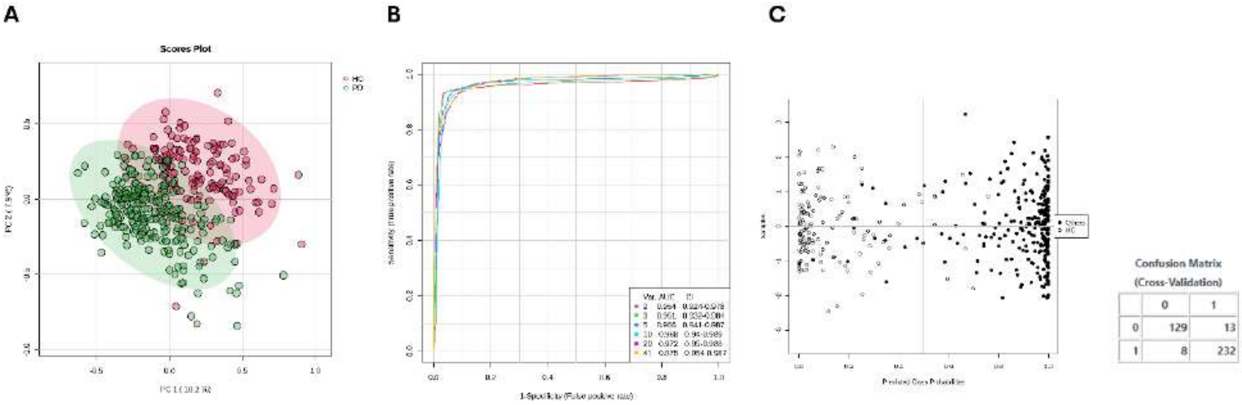


**Figure S2. Validation methods for clustering models analysing serum profiles of controls and Parkinson's disease patients, using supervised algorithms, AUC curves, and support vector machines (SVMs). A,** Score plots from the PCA conducted on serum metabolomic profiles of PD patients compared HC (PERMANOVA validation: F= 21.39, R = 0.64, p = 0.001). **B,** PLS-DA models were validated using the area under the ROC curve, with the x-axis representing the false positive rate and the y-axis representing the true positive rate across the entire cohort of Parkinson's Disease and Healthy Control MVA models, respectively. **C,** Support Vector Machine results used to assess the model's discriminative performance, including the corresponding confusion matrix and the number of misclassified samples.

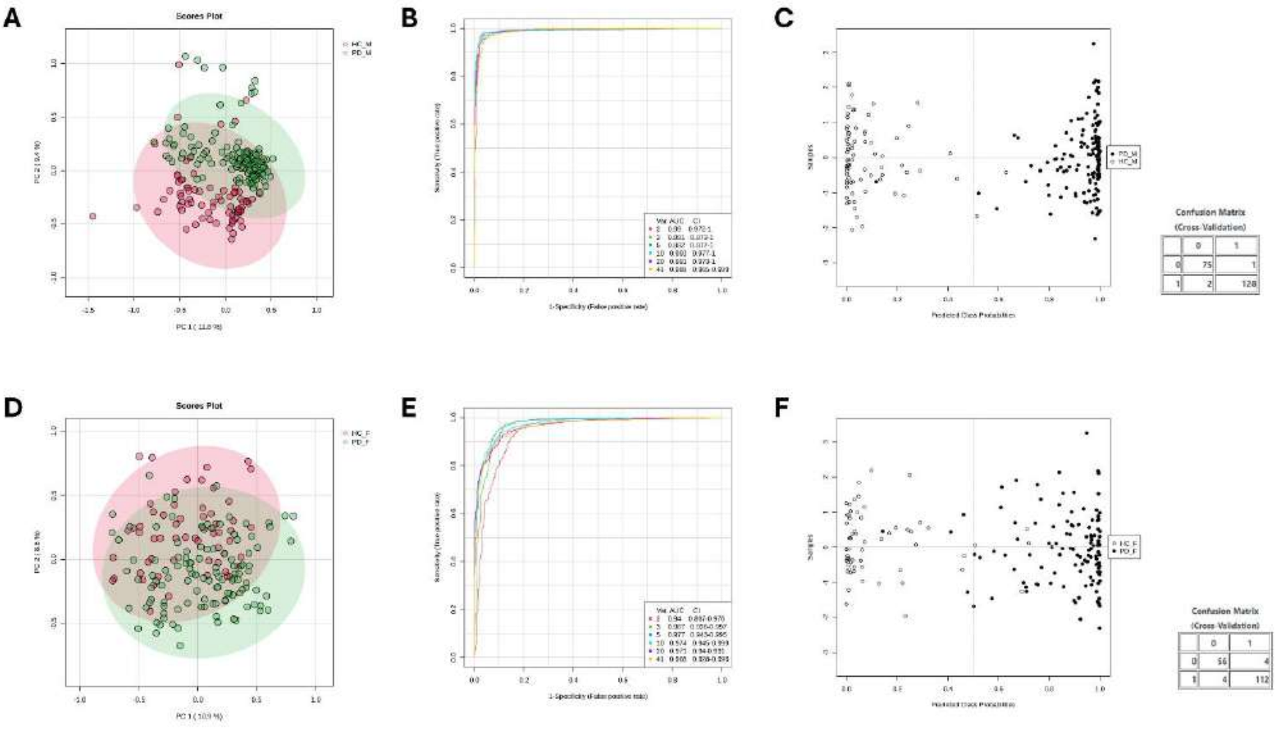


**Figure S3. Validation methods for clustering models analysing serum profiles of controls and Parkinson's disease patients, stratified by sex, using supervised algorithms, AUC curves, and support vector machines (SVMs).** Score plots from the PCA conducted on serum metabolomic profiles of PD patients compared to sex matched HC (**panel A**, Male cohort; PERMANOVA validation: F = 31.53, R = 0.66, p = 0.001; **panel D**, Female cohort, PERMANOVA validation: F = 22.35, R = 0.11, p = 0.001). **Panels B, E,** illustrate the PLS-DA models validated through the area under the ROC curve, where the x-axis represents the false positive rate and the y-axis the true positive rate in males and females' MVA models, respectively. **Panels C, F,** Support Vector Machine results used to assess the discriminative performance of the model, including the corresponding confusion matrix and the number of misclassified samples.

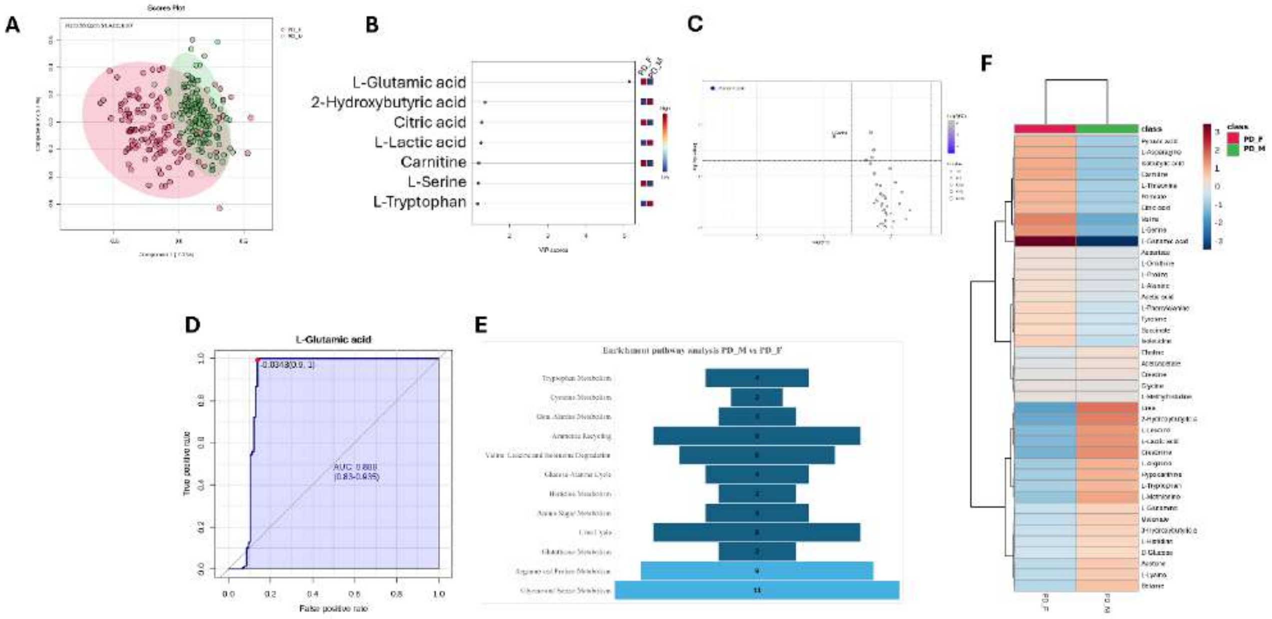


**Figure S4. NMR metabolomics identifies distinct serum metabolomic profiles between male and female patients with PD.** **A**, PLS-DA score plot is shown in Cartesian coordinates, with the percentage of variance explained by the primary (PC1: 7.1) and secondary (PC2: 5.2) components displayed along the x and y axes, respectively. The supervised model was constructed based on serum metabolite concentrations from patients with idiopathic Parkinson's disease, with 129 males and 115 females. The validation index was determined using 10-fold cross-validation, with metrics such as R2, Q2, and accuracy reported at the top of the plots. **B**, Variable Importance in Projection (VIP) scores identifying metabolites contributing to group separation; only metabolites with VIP > 1 were considered significant. **C,** Robust volcano plots highlighting upregulated (red) and downregulated (blue) metabolites in PD patients relative to males, using a fold-change threshold of 1.5 and a p-value < 0.05. **D**, Receiver Operating Characteristic (ROC) curves evaluating biomarker performance, where the x-axis represents the false-positive rate and the y-axis the true-positive rate. Each ROC curve includes the empirical ROC curve and the chance diagonal (45° line from (0, 0) to (1, 1)). **E**, Pathway enrichment analyses based on 1H-NMR metabolomics data from male compared to female PD patients compared with their respective HCs. Bars represent the number of detected metabolites (hits) mapped to each pathway. Pathways were considered significant when Hits ≥ 2, p < 0.05, Holm-adjusted p < 0.05, and FDR < 0.05. Darker colours indicate lower p-values. Analyses were performed using the Small Molecule Pathway Database (SMPDB) with Homo sapiens selected as the reference organism. **F**, The heatmap analysis highlights metabolic changes showing upregulated metabolites in red and downregulated in blue, respectively.

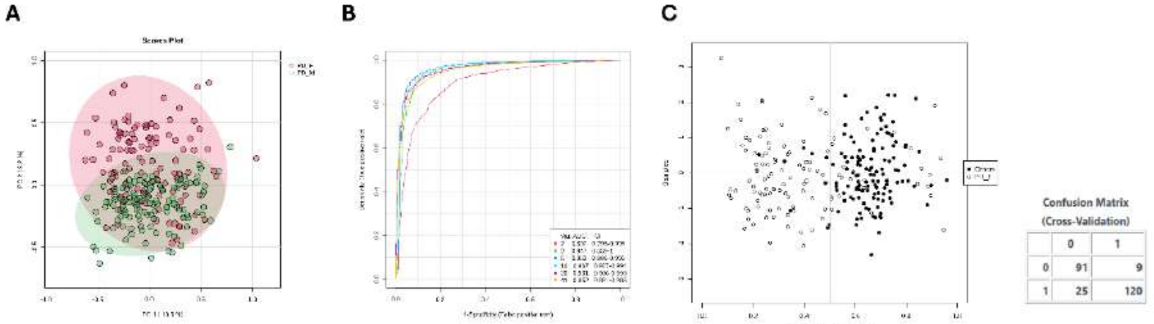


**Figure S5. Validation approaches for clustering models compare serum profiles of Parkinson's disease patients within the whole cohort, differentiated by gender, utilizing supervised algorithms, AUC curves, and support vector machines (SVMs). A,** Score plots from the PCA conducted on serum metabolomic profiles of PD patients compared within sex (PERMANOVA validation: F = 12.25, R = 0.36, p = 0.003). **B,** PLS-DA models validated through the area under the ROC curve, where the x-axis represents the false positive rate and the y-axis the true positive rate in males' and females' MVA models, respectively. **C,** Support Vector Machine results used to assess the discriminative performance of the model, including the corresponding confusion matrix and the number of misclassified samples.

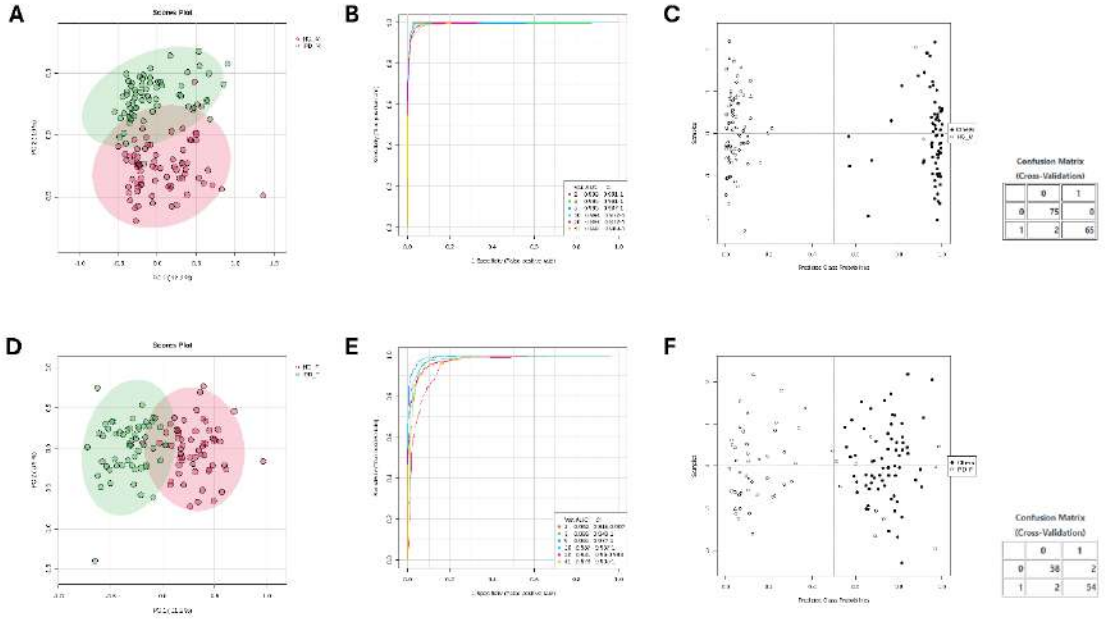


**Figure S6. Validation methods for clustering models analysing serum profiles of controls and idiopathic Parkinson's disease patients, stratified by sex, using supervised algorithms, AUC curves, and support vector machines (SVMs).** Score plots from the PCA conducted on serum metabolomic profiles of iPD patients compared to sex matched HC (**panel A**, Male cohort; PERMANOVA validation: F = 67.78, R = 0.33, p = 0.001; **panel D**, Female cohort, PERMANOVA validation: F = 78.76, R = 0.40, p = 0.001). **Panels B, E,** illustrate the PLS-DA models validated through the area under the ROC curve, where the x-axis represents the false positive rate and the y-axis the true positive rate in males' and females' MVA models, respectively. **Panels C, F,** Support Vector Machine results used to assess the discriminative performance of the model, including the corresponding confusion matrix and the number of misclassified samples.

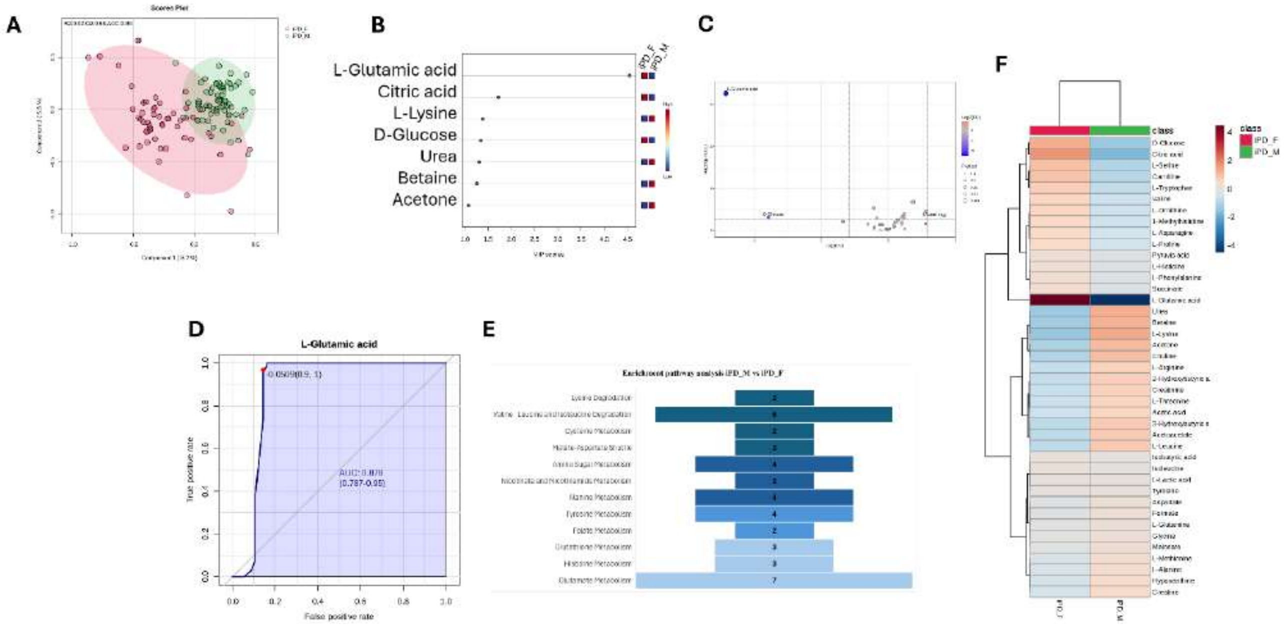


**Figure S7. NMR metabolomics identifies distinct serum metabolomic profiles between male and female patients with iPD.** **A**, PLS-DA score plot is shown in Cartesian coordinates, with the percentage of variance explained by the primary (PC1: 8.2) and secondary (PC2: 5.8) components displayed along the x and y axes, respectively. The supervised model was constructed based on serum metabolite concentrations from patients with idiopathic Parkinson's disease, with 65 males and 56 females. The validation index was determined using 10-fold cross-validation, with metrics such as R2, Q2, and accuracy reported at the top of the plots. **B**, Variable Importance in Projection (VIP) scores identifying metabolites contributing to group separation; only metabolites with VIP > 1 were considered significant. **C,** Robust volcano plots highlighting upregulated (red) and downregulated (blue) metabolites in iPD patients relative to males, using a fold-change threshold of 1.5 and a p-value < 0.05. **D**, Receiver Operating Characteristic (ROC) curves evaluating biomarker performance, where the x-axis represents the false-positive rate and the y-axis the true-positive rate. Each ROC curve includes the empirical ROC curve and the chance diagonal (45° line from (0, 0) to (1, 1)). **E**, Pathway enrichment analyses based on 1H-NMR metabolomics data from male compared to female iPD patients compared with their respective HCs. Bars represent the number of detected metabolites (hits) mapped to each pathway. Pathways were considered significant when Hits ≥ 2, $p < 0.05$, Holm-adjusted $p < 0.05$, and FDR < 0.05. Darker colours indicate lower p-values. Analyses were performed using the Small Molecule Pathway Database (SMPDB) with Homo sapiens selected as the reference organism. **F**, The heatmap analysis highlights metabolic changes showing upregulated metabolites in red and downregulated in blue, respectively.

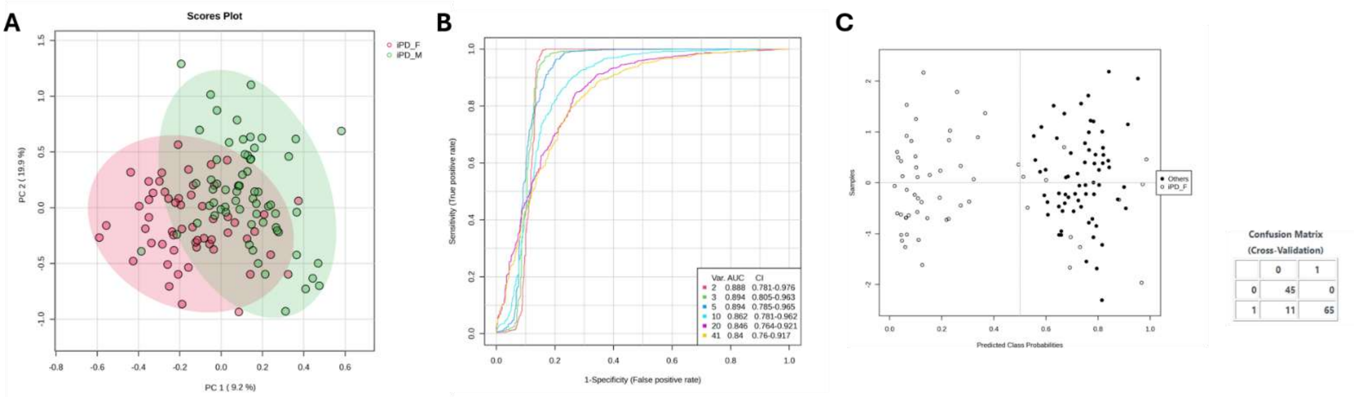


**Figure S8. Validation methods for clustering models analysing serum profiles of male and female idiopathic Parkinson's disease patients, using supervised algorithms, AUC curves, and support vector machines (SVMs).** Score plots from the PCA conducted on serum metabolomic profiles of iPD patients stratified by gender (**PERMANOVA** validation: F = 68.42, R = 0.33, p = 0.001; **B** illustrate the PLS-DA models validated through the area under the ROC curve, where the x-axis represents the false positive rate and the y-axis the true positive rate in males' and females' MVA models, respectively. **C** Support Vector Machine results used to assess the model's discriminative performance, including the corresponding confusion matrix and the number of misclassified samples.

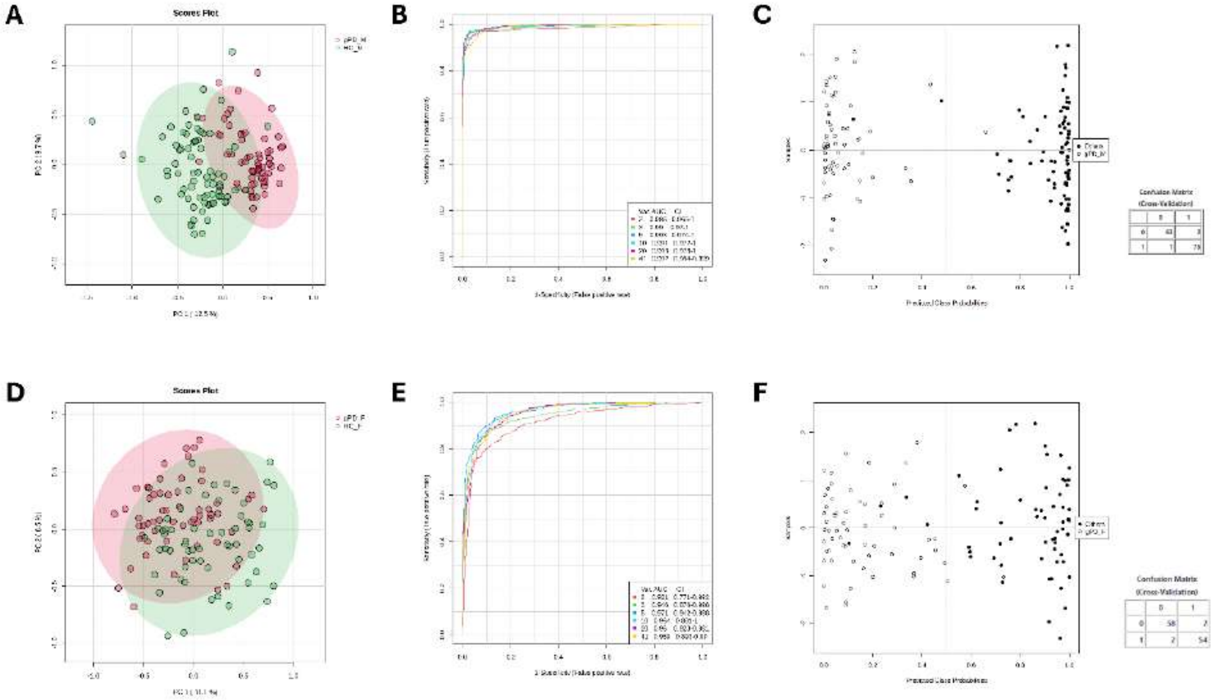


**Figure S9. Validation methods for clustering models analysing serum profiles of controls and genetic Parkinson's disease patients, stratified by sex, using supervised algorithms, AUC curves, and support vector machines (SVMs).** Score plots from the PCA conducted on serum metabolomic profiles of gPD patients compared to sex matched HC (**panel A**, Male cohort; PERMANOVA validation: F = 68.42, R = 0.33, p = 0.001,; **panel D**, Female cohort, PERMANOVA validation: F = 18.76, R = 0.20, p = 0.001). **Panels B, E** illustrate the PLS-DA models validated through the area under the ROC curve, where the x-axis represents the false positive rate and the y-axis the true positive rate in males' and females' MVA models, respectively. **Panels C, F,** Support Vector Machine results used to assess the discriminative performance of the model, including the corresponding confusion matrix and the number of misclassified samples.

**Table S1. Demographic and clinical characteristics compared between idiopathic PD and genetic PD enrolled in the serum collection for NMR analysis.**

| Sex | Clinical characteristics | Idiopathic PD | | | | Genetic PD | | | | *p* value$^a$ |
|---|---|---|---|---|---|---|---|---|---|---|
| | | N | Median | IQR | | N | Median | IQR | | |
| Male | Age (years) | 65 | 67.0 | 61.0 | 74.0 | 64 | 68.0 | 61.5 | 72.0 | 0.938 |
| | Age at onset (years) | 65 | 61.0 | 55.0 | 66.0 | 64 | 60.0 | 55.5 | 65.5 | 0.869 |
| | Disease duration (years) | 65 | 5.0 | 3.0 | 8.0 | 64 | 6.0 | 3.0 | 9.5 | 0.610 |
| | LEDD (mg/die) | 65 | 400.0 | 300.0 | 560.0 | 64 | 451.0 | 320.0 | 724.0 | 0.304 |
| | MDS-UPDRS III | 65 | 24.0 | 13.0 | 35.0 | 64 | 22.5 | 14.5 | 31.0 | 0.668 |
| Female | Age (years) | 56 | 69.5 | 62.5 | 74.0 | 60 | 68.0 | 63.5 | 73.5 | 0.560 |
| | Age at onset (years) | 56 | 64.0 | 56.5 | 69.0 | 60 | 61.5 | 52.5 | 65.0 | **0.042** |
| | Disease duration (years) | 56 | 5.0 | 3.0 | 7.0 | 60 | 7.0 | 3.5 | 12.0 | **0.012** |
| | LEDD (mg/die) | 56 | 400.0 | 302.5 | 600.0 | 60 | 483.0 | 300.0 | 645.0 | 0.730 |
| | MDS-UPDRS III | 56 | 21.0 | 13.0 | 28.5 | 60 | 17.5 | 12.0 | 30.0 | 0.525 |

Abbreviations: N, number of subjects; IQR, interquartile range; LEDD, Levodopa equivalent daily dose; MDS-UPDRS III, Movement Disorders Society Unified Parkinson's Disease Rating Scale, part III. [a]Mann-Whitney test

**Table S2. Demographic and clinical characteristics compared between male and female in both idiopathic PD and genetic PD patients enrolled in the serum collection for NMR analysis.**

| PD subtype | Clinical characteristics | Male | | | | Female | | | | *p* value[a] |
|---|---|---|---|---|---|---|---|---|---|---|
| | | N | Median | IQR | | N | Median | IQR | | |
| Idiopathic PD | Age (years) | 65 | 67.0 | 61.0 | 74.0 | 56 | 69.5 | 62.5 | 74.0 | 0.321 |
| | Age at onset (years) | 65 | 61.0 | 55.0 | 66.0 | 56 | 64.0 | 56.5 | 69.0 | 0.196 |
| | LEDD (mg/die) | 65 | 400.0 | 300.0 | 560.0 | 56 | 400.0 | 302.5 | 600.0 | 0.402 |
| | Disease duration (years) | 65 | 5.0 | 3.0 | 8.0 | 56 | 5.0 | 3.0 | 7.0 | 0.532 |
| | MDS-UPDRS III | 65 | 24.0 | 13.0 | 35.0 | 56 | 21.0 | 13.0 | 28.5 | 0.191 |
| Genetic PD | Age (years) | 64 | 68.0 | 61.5 | 72.0 | 60 | 68.0 | 63.5 | 73.5 | 0.676 |
| | Age at onset (years) | 64 | 60.0 | 55.5 | 65.5 | 60 | 61.5 | 52.5 | 65.0 | 0.556 |
| | LEDD (mg/die) | 64 | 451.0 | 320.0 | 724.0 | 60 | 483.0 | 300.0 | 645.0 | 0.795 |
| | Disease duration (years) | 64 | 6.0 | 3.0 | 9.5 | 60 | 7.0 | 3.5 | 12.0 | 0.151 |
| | MDS-UPDRS III | 64 | 22.5 | 14.5 | 31.0 | 60 | 17.5 | 12.0 | 30.0 | 0.167 |

Abbreviations: N, number of subjects; IQR, interquartile range; LEDD, Levodopa equivalent daily dose; MDS-UPDRS III, Movement Disorders Society Unified Parkinson's Disease Rating Scale, part III. [a]Mann-Whitney test

**Table S3. List of pathogenic mutations in *LRRK2, GBA1, PARK2, PINK1, PARK7* and *TMEM175* in genetic-PD subtype**

| CHR | Genomic position (hg38) | dbSNP | Gene | RefSEq | Nucleotide Change | AA Change | Exonic Function | Cl | MAF gnomAD v 4.1 | CADD phred | PD patients N |
|---|---|---|---|---|---|---|---|---|---|---|---|
| 12 | 40340400 | rs34637584 | LRRK2 | NM_198578 | c.G6055A | p.G2019S | NSV | P | 0.0002721 | 35 | 14 |
| 12 | 40310434 | rs33939927 | LRRK2 | NM_198578 | c.C4321T | p.R1441C | NSV | P | 0.0000195 | 26.7 | 2 |
| 12 | 39859182 | rs34594498 | LRRK2 | NM_198578 | c.C1256T | p.A419V | NSV | P | 0.0001493 | 24.9 | 1 |
| 1 | 155266052 | rs76763715 | GBA1 | NM_000157 | c.A1226G | p.N409S | NSV | P | 0.001728 | 24.1 | 9 |
| 1 | 155266585 | rs2230288 | GBA1 | NM_000157 | c.G1093A | p.E365K | NSV | P | 0.01338 | 16.1 | 7 |
| 1 | 155296664 | rs75548401 | GBA1 | NM_000157 | c.C1223T | p.T408M | NSV | P | 0.008175 | 21.3 | 3 |
| 1 | 155265461 | rs421016 | GBA1 | NM_000157 | c.T1448C | p.L483P | NSV | P | 0.00007547 | 24.7 | 2 |
| 1 | 155267667 | rs367968666 | GBA1 | NM_000157 | c.T882G | p.H294Q | NSV | P | 0.0001678 | 12.25 | 2 |
| 1 | 155270905 | rs139626710 | GBA1 | NM_000157 | c.A49G | p.R17G | NSV | P | 0.00002119 | 8.03 | 1 |
| 1 | 155270204 | NA | GBA1 | NM_000157 | c.C198A | p.D66E | NSV | P | NA | 6.32 | 1 |
| 1 | 155268415 | rs381427 | GBA1 | NM_000157 | c.T689A | p.V230E | NSV | P | 0.00000339 | 22.3 | 1 |
| 1 | 155268386 | NA | GBA1 | NM_000157 | c.C718T | p.P240S | NSV | P | NA | 22.4 | 1 |
| 1 | 155266579 | NA | GBA1 | NM_000157 | c.C1099T | p.H367Y | NSV | P | NA | 25.7 | 1 |
| 1 | 155266470 | rs121908307 | GBA1 | NM_000157 | c.G1208C | p.S403T | NSV | P | 8.476e-7 | 22.8 | 1 |
| 1 | 155265999 | rs149171124 | GBA1 | NM_000157 | c.G1279A | p.E427K | NSV | P | 0.0002229 | 19.29 | 1 |
| 1 | 155265936 | rs1064651 | GBA1 | NM_000157 | c.G1342C | p.D448H | NSV | P | 0.0001043 | 23.3 | 1 |
| 1 | 155265414 | rs369068553 | GBA1 | NM_000157 | c.G1495C | p.V499L | NSV | P | 0.00004747 | 17.25 | 1 |
| 1 | 155265300 | NA | GBA1 | NM_000157 | c.G1515T | p.K505N | NSV | P | NA | 19.17 | 1 |
| 6 | 160929107 | rs775091228 | PRKN | NM_004562 | c.G1358A | p.W453X | Stop_gain | P | 0.00001186 | 44 | 2 |
| 6 | 161364788 | rs34424986 | PRKN | NM_004562 | c.C823T | p.R275W | NSV | P | 0.003732 | 26.1 | 2 |
| 6 | 160939137 | rs55830907 | PRKN | NM_004562 | c.C1204T | p.R402C | NSV | P | 0.001937 | 25.1 | 4 |
| 6 | 161841660 | rs55774500 | PRKN | NM_004562 | c.C245A | p.A82E | NSV | P | 0.002815 | 3.14 | 5 |
| 6 | 161633103 | rs9456735 | PRKN | NM_004562 | c.A574C | p.M192L | NSV | P | 0.0001356 | 20.7 | 1 |
| 6 | 161552303 | rs144032774 | PRKN | NM_004562 | c.G701A | p.R234Q | NSV | P | 0.0001280 | 20.9 | 1 |
| 6 | 161552285 | rs137853054 | PRKN | NM_004562 | c.C719T | p.T240M | NSV | P | 0.0001802 | 23.4 | 1 |
| 6 | 161552274 | NA | PRKN | NM_004562 | c.G730C | p.V244L | NSV | P | NA | 6.93 | 1 |
| 6 | 160939116 | NA | PRKN | NM_004562 | c.G1225T | p.E409X | Stop_gain | P | NA | 40 | 1 |

| | | | | | | | | | | | |
|---|---|---|---|---|---|---|---|---|---|---|---|
| 6 | 160939097 | rs778125254 | PRKN | NM_004562 | c.C1244A | p.T415N | NSV | P | 0.00000339 | 25.2 | 1 |
| 1 | 20311548 | rs138302371 | PINK1 | NM_032409 | c.C587T | p.P196L | NSV | P | 0.0003610 | 17.46 | 2 |
| 1 | 20322561 | rs74315356 | PINK1 | NM_032409 | c.G1311A | p.W437X | Stop_gain | P | 0.00000677 | 47 | 2 |
| 1 | 20311463 | rs768091663 | PINK1 | NM_032409 | c.G502C | p.A168P | NSV | P | 0.00000847 | 22.2 | 1 |
| 1 | 20311519 | rs143204084 | PINK1 | NM_032409 | c.G558C | p.K186N | NSV | P | 0.0002568 | 14.84 | 1 |
| 1 | 20318092 | NA | PINK1 | NM_032409 | c.C872A | p.A291D | NSV | P | NA | 26.3 | 1 |
| 1 | 20319083 | rs376323248 | PINK1 | NM_032409 | c.C976T | p.R326C | NSV | P | 0.00000508 | 26.9 | 1 |
| 1 | 20324025 | rs531477772 | PINK1 | NM_032409 | c.G1573A | p.D525N | NSV | P | 0.00009152 | 23.3 | 1 |
| 1 | 7910874 | rs71653619 | PARK7 | NM_007262 | c.G293A | p.R98Q | NSV | RF | 0.01043 | 20.9 | 14 |
| 1 | 7909346 | rs781094807 | PARK7 | NM_007262 | c.252+2->A | | splicing | N | 0.0002733 | NA | 2 |
| 4 | 958630 | rs752406714 | TMEM175 | NM_032326 | c.430_431del | p.V147Dfs*104 | Fs_del | P | 0.00000256 | NA | 4 |
| 4 | 964474 | rs565504915 | TMEM175 | NM_032326 | c.1281_1282del | p.A429Qfs*120 | Fs_del | P | 0.0003095 | NA | 3 |
| 4 | 958646 | NA | TMEM175 | NM_032326 | c.446_462del | p.A149Gfs*97 | Fs_del | P | NA | NA | 3 |
| 4 | 954054 | rs542936413 | TMEM175 | NM_032326 | c.C103T | p.R35C | NSV | P | 0.00002542 | 28.7 | 3 |
| 4 | 964406 | rs75307864 | TMEM175 | NM_032326 | c.C1213G | p.L405V | NSV | P | 0.003832 | 23.3 | 4 |
| 4 | 964433 | rs140597786 | TMEM175 | NM_032326 | c.C1240T | p.R414W | NSV | P | 0.004833 | 23.2 | 3 |
| 4 | 964197 | rs147762522 | TMEM175 | NM_032326 | c.G1004A | p.R335H | NSV | P | 0.001076 | 24.6 | 3 |
| 4 | 956673 | rs142778595 | TMEM175 | NM_032326 | c.T233C | p.I78T | NSV | P | 0.00005424 | 23.4 | 2 |
| 4 | 957441 | rs200834686 | TMEM175 | NM_032326 | c.A313G | p.T105A | NSV | P | 0.0001136 | 22.21 | 2 |
| 4 | 962038 | rs746980739 | TMEM175 | NM_032326 | c.C778T | p.R260C | NSV | P | 0.00004068 | 25.1 | 1 |
| 4 | 962068 | rs750645874 | TMEM175 | NM_032326 | c.G808A | p.A270T | NSV | P | 0.00000762 | 25.0 | 1 |
| 4 | 962100 | NA | TMEM175 | NM_032326 | c.C840G | p.I280M | NSV | P | NA | 24.7 | 1 |
| 4 | 964050 | rs778399444 | TMEM175 | NM_032326 | c.C857T | p.P286L | NSV | P | 0.00000254 | 23.9 | 1 |
| 4 | 964170 | rs148627215 | TMEM175 | NM_032326 | c.C977T | p.A326V | NSV | P | 0.00000169 | 7.41 | 1 |
| 4 | 964236 | rs147975675 | TMEM175 | NM_032326 | c.C1043T | p.S348L | NSV | P | 0.00005171 | 24.5 | 1 |
| 4 | 964464 | rs142744759 | TMEM175 | NM_032326 | c.C1271T | p.A424V | NSV | P | 0.00001188 | 22.6 | 1 |
| 4 | 964634 | rs201314478 | TMEM175 | NM_032326 | c.C1441T | p.R481W | NSV | P | 0.002678 | 10.6 | 1 |

Abbreviations: CHR, Chromosome; hg38, human genome assembly GRCh38; dbSNP, reference number in Single Nucleotide Polymorphism (SNP) database; ref seq, reference number of the gene transcript; AA Change, amino acid change; CI, clinical interpretation; P, Pathogenic; RF, risk factor; N, novel; PD, Parkinson's disease; CADD phred, Combined Annotation Dependent Depletion; Fs_del, frameshift deletion; NSV, non-synonymous variant; MAF, Minor Allele Frequency, was referred to gnomAD v4.1 database; NA, Not Annotat

**Table S4. The Out-Of-Bag (OOB) error was determined using a random forest method to classify clustering patterns among metabolomic profiles of Parkinson's Disease (PD), stratified by gender and genotypes, and compared with sex and age-matched healthy controls.** The table summarises the performance of Random Forest classification models applied to the entire cohort, comprising idiopathic and genetic PD patients, stratified by sex. For each model, the class error (class.error) and the OOB error are reported, with the latter serving as an internal estimate of the model's accuracy. The models showed very low OOB errors (0–0.016) across all comparisons.

| | HC | PD | OOB error | Total OOB |
|---|---|---|---|---|
| HC | 134 | 3 | 0.0219 | 0.007 |
| PD | 0 | 245 | 0.0 | |
| | **HC_M** | **PD_M** | **OOB error** | **Total OOB** |
| HC_M | 76 | 1 | 0.013 | 0.004 |
| PD_M | 0 | 129 | 0 | |
| | **HC_F** | **PD_F** | **OOB error** | **Total OOB** |
| HC_F | 59 | 1 | 0.016 | 0.005 |
| PD_F | 0 | 116 | 0 | |
| | **HC_M** | **iPD_M** | **OOB error** | **Total OOB** |
| HC_M | 77 | 0 | 0 | 0 |
| iPD_M | 0 | 65 | 0 | |
| | **HC_F** | **iPD_F** | **OOB error** | **Total OOB** |
| HC_F | 60 | 0 | 0 | 0.008 |
| iPD_F | 1 | 55 | 0.01 | |
| | **HC_M** | **gPD_M** | **OOB error** | **Total OOB** |
| HC_M | 77 | 0 | 0 | 0 |
| gPD_M | 0 | 64 | 0 | |
| | **HC_F** | **gPD_F** | **OOB error** | **Total OOB** |
| HC_F | 59 | 1 | 0.016 | 0.016 |
| gPD_F | 1 | 59 | 0.016 | |

**Table S5. Robust Volcano plot results for PD patients compared with HC.**

| Metabolites PD vs HC | FC | log2(FC) | raw.pval | -log10(p) |
|---|---|---|---|---|
| L-Threonine | 2.5193 | 1.333 | 4.54E-19 | 18.343 |
| Acetic acid | 0.46146 | -1.1157 | 1.39E-15 | 14.858 |
| Choline | 0.58187 | -0.78123 | 4.65E-14 | 13.333 |
| Pyruvic acid | 0.5729 | -0.80365 | 7.33E-10 | 9.1349 |
| L-Methionine | 1.631 | 0.70578 | 3.49E-09 | 8.4566 |
| Malonate | 0.63602 | -0.65285 | 5.75E-05 | 4.2402 |

Results from univariate analysis were presented using a Robust Volcano plot of metabolite concentrations detected in the 1d-CPMG NMR spectrum. The analysis combines the Fold change (FC) test, calculated as the ratio of average concentrations between the PD/HC clusters —using a threshold of 1.5—and the p-value from the T-Test, which is considered significant if less than 0.05.

**Table S6. Pathway enrichment analysis across PD patients compared with HC**

| Dysregulated Pathways | Hits | Raw p | Holm p | FDR | Metabolites |
|---|---|---|---|---|---|
| Aspartate Metabolism | 6 | 1.65E-07 | 1.12E-05 | 3.84E-06 | Acetic acid; L-Glutamic acid; Asparagine; Aspartate; L-Glutamine; Arginine. |
| Butyrate Metabolism | 2 | 2.51E-07 | 1.68E-06 | 2.39E-06 | Succinic acid; Acetoacetic acid. |
| Oxidation of Branched Chain Fatty Acids | 2 | 3.70E-07 | 2.44E-05 | 2.18E-06 | Succinic acid; Carnitine. |
| Ketone Body Metabolism | 4 | 5.28E-07 | 3.43E-05 | 3.16E-06 | Succinic acid; Carnitine; Acetoacetic acid; 3-Hydroxybutyrate. |
| Citric Acid Cycle | 3 | 1.22E-04 | 7.78E-03 | 1.22E-03 | Succinic acid; Citric acid; Pyruvic acid. |
| Fatty Acid Biosynthesis | 3 | 1.87E-03 | 1.18E-02 | 1.64E-02 | Acetic acid; Acetoacetic acid; 3-Hydroxybutyrate. |
| Carnitine Synthesis | 4 | 1.14E-02 | 1.95E-02 | 2.36E-02 | Succinic acid; Carnitine; Glycine; Lysine. |
| Amino Sugar Metabolism | 4 | 1.16E-02 | 1.95E-02 | 2.36E-02 | Acetic acid; L-Glutamine; L-Glutamic acid; Pyruvic acid. |
| Arginine and Proline Metabolism | 9 | 1.81E-02 | 3.43E-02 | 3.39E-02 | L-Glutamic acid; Proline; Aspartate; Succinic acid; Urea; L-Arginine; L-Ornithine; Creatine; Glycine. |
| Glutamic acid Metabolism | 7 | 1.09E-02 | 1.35E-02 | 3.84E-02 | Glutamic acid; Pyruvic acid; Aspartate; Glycine; Alanine; L-Glutamine; Succinic acid. |
| Betaine Metabolism | 3 | 1.17E-02 | 1.66E-02 | 3.84E-02 | Betaine; Choline; Methionine. |
| Valine Leucine and Isoleucine Degradation | 6 | 2.19E-02 | 3.23E-02 | 3.02E-02 | Acetoacetic acid; L-Glutamic acid; Isoleucine; Succinic acid; L-Valine; L-Leucine. |
| Warburg Effect | 7 | 3.00E-02 | 3.65E-02 | 3.31E-02 | Citric acid; D-Glucose; L-Glutamic acid; Pyruvic acid; Succinic acid; Lactic acid; L-Glutamine. |
| Urea Cycle | 8 | 1.33E-02 | 3.20E-02 | 3.89E-02 | L-Glutamic acid; L-Alanine; L-Aspartic acid; L-Ornithine; Pyruvic acid; Urea; L-Arginine; L-Glutamine. |

List of enriched pathways resulting from the comparison between PD patients' serum profiles and those of HC. The analysis was performed using the Small Molecule Pathways Database (SMPDB) with *Homo sapiens* selected as the organism. Hits refer to the number of metabolites involved in the pathways, as listed in the ‘Metabolites’ column. The pathways were deemed statistically significant if they had Hits $\geqslant 2$, a p-value $< 0.05$, an adjusted p-value from the Holm-Bonferroni test (Holm p), and a False Discovery Rate (FDR) $< 0.05$.

**Table S7. Robust Volcano plot results related to PD patients compared to HC stratified by gender**

| Metabolites PD_M vs HC_M | FC | log2(FC) | raw.pval | -log10(p) |
|---|---|---|---|---|
| L-Glutamic acid | 0.39313 | -1.3469 | 3.32E-18 | 17.479 |
| L-Tryptophan | 2.729 | 1.4483 | 2.04E-12 | 11.691 |
| Acetic acid | 0.42769 | -1.2254 | 3.15E-08 | 7.5016 |
| L-Methionine | 1.7157 | 0.77879 | 1.29E-07 | 6.89 |
| Pyruvic acid | 0.52837 | -0.92038 | 1.50E-06 | 5.8251 |
| Choline | 0.62448 | -0.67928 | 1.30E-05 | 4.8877 |
| Malonate | 0.61039 | -0.71219 | 1.54E-05 | 4.8115 |
| **Metabolites PD_F vs HC_F** | **FC** | **log2(FC)** | **raw.pval** | **-log10(p)** |
| Choline | 0.53697 | -0.89708 | 5.78E-11 | 10.238 |
| Acetic acid | 0.5019 | -0.99453 | 5.03E-09 | 8.2983 |
| L-Threonine | 2.2907 | 1.1958 | 5.24E-08 | 7.2809 |
| Pyruvic acid | 0.61672 | -0.69732 | 0.000105 | 3.9789 |
| 3-Hydroxybutyric acid | 1.6014 | 0.67934 | 0.0008 | 3.0968 |

Results from univariate analysis were presented using a Robust Volcano plot of metabolite concentrations detected in the 1d-CPMG NMR spectrum. The analysis combines the Fold change (FC) test, calculated as the ratio of average concentrations between the PD/HC clusters stratified by sex—using a threshold of 1.5—and the p-value from the T-Test, which is considered significant if less than 0.05

**Table S8. Pathway enrichment analysis across male and female PD patients compared with HC**

| Dysregulated Pathways PD_M | Hits | Raw p | Holm p | FDR | Metabolites |
|---|---|---|---|---|---|
| Glutamic acid Metabolism | 7 | 1.35E-10 | 5.93E-04 | 3.49E-04 | L-Glutamic acid; Pyruvic acid; Aspartate; Glycine; L-Alanine; L-Glutamine; Succinic acid. |
| Glycolysis | 2 | 1.59E-08 | 6.34E-07 | 3.58E-05 | Pyruvic acid;Glucose. |
| Glutathione Metabolism | 3 | 4.25E-08 | 2.98E-08 | 2.98E-08 | Glycine; L-Glutamic acid; L-Ornithine. |
| Phenylalanine and Tyrosine Metabolism | 4 | 7.77E-08 | 5.36E-08 | 2.72E-08 | Acetoacetic acid; L-Glutamic acid; L-Tyrosine; L-Phenylalanine. |
| Histidine Metabolism | 3 | 3.01E-07 | 2.01E-07 | 5.26E-07 | L-Glutamic acid; L-Histidine; 1-Methylhistidine. |
| Tyrosine Metabolism | 4 | 2.55E-05 | 1.66E-05 | 2.98E-05 | Acetoacetic acid; L-Glutamic acid; L-Tyrosine; L-Phenylalanine. |
| Tryptophan Metabolism | 4 | 3.24E-05 | 2.01E-05 | 2.52E-05 | L-Glutamic acid; L-Tryptophan; Formic acid; L-Alanine. |
| Beta-Alanine Metabolism | 3 | 4.19E-05 | 2.68E-05 | 3.73E-05 | L-Glutamic acid;Aspartate; L-Histidine. |
| Malate-Aspartate Shuttle | 2 | 4.26E-05 | 2.69E-05 | 3.73E-05 | Aspartate; L-Glutamic acid. |
| Amino Sugar Metabolism | 4 | 1.01E-03 | 5.83E-03 | 5.41E-03 | L-Glutamic acid; Pyruvic acid; Acetoacetic acid; L-Glutamine. |
| Ammonia Recycling | 8 | 1.02E-03 | 6.24E-03 | 7.16E-03 | L-Glutamic acid; Pyruvic acid; L-Asparagine; Glycine; L-Serine; Aspartate; L-Histidine; L-Glutamine. |
| Lysine Degradation | 2 | 1.22E-03 | 7.29E-03 | 7.73E-03 | L-Glutamic acid; L-Lysine. |
| Aspartate Metabolism | 6 | 8.87E-03 | 5.23E-03 | 5.17E-03 | L-Glutamic acid; Acetic acid; L-Asparagine; Aspartate; L-Glutamine; L-Arginine. |
| Glycine and Serine Metabolism | 11 | 2.05E-02 | 1.17E-02 | 1.03E-02 | Betaine, Creatine, Glycine, L-Glutamic acid, L-Alanine, L-Threonine, L-Serine, L-Ornithine, Pyruvic acid, L-Methionine, L-Arginine. |
| Cysteine Metabolism | 2 | 2.07E-02 | 1.14E-02 | 3.70E-02 | L-Glutamic acid, Pyruvic acid. |
| **Dysregulated Pathways PD_F** | **Hits** | **Raw p** | **Holm p** | **FDR** | **Metabolites** |
| Betaine Metabolism | 3 | 7.92E-04 | 4.75E-09 | 5.04E-09 | Betaine; Choline; Methionine |
| Fatty Acid Biosynthesis | 3 | 1.18E-03 | 4.17E-02 | 4.23E-02 | Acetic acid, Acetoacetic acid, 3-Hydroxybutyrate. |
| Glycine and Serine Metabolism | 11 | 1.30E-03 | 3.13E-02 | 4.13E-02 | Betaine, Creatine, Glycine, L-Glutamic acid, L-Alanine, L-Threonine, L-Serine, L-Ornithine, Pyruvic acid, L-Methionine, L-Arginine. |
| Spermidine and Spermine Biosynthesis | 2 | 1.17E-02 | 4.34E-02 | 4.02E-02 | L-Ornithine; L-Methionine. |

| | | | | | |
|---|---|---|---|---|---|
| Urea Cycle | 8 | 1.36E-02 | 3.10E-02 | 2.38E-02 | L-Glutamic acid; L-Alanine; Aspartate; L-Ornithine; Pyruvic acid; Urea; Arginine; L-Glutamine. |
| Phosphatidylethanolamine Biosynthesis | 2 | 2.20E-02 | 1.52E-02 | 2.72E-02 | Choline; L-Serine. |
| Methionine Metabolism | 5 | 2.92E-02 | 1.87E-02 | 2.92E-02 | Betaine,Glycine; L-Serine; L-Methionine; Choline. |
| Amino Sugar Metabolism | 4 | 2.04E-02 | 3.75E-02 | 4.7E-02 | L-Glutamic acid; L-Glutamine; Pyruvic acid; Acetic acid. |

List of enriched pathways resulted from the comparison of PD patients' serum profile stratified for sex and compared to matched HC. The analysis was performed using the Small Molecule Pathways Database (SMPDB) with *Homo sapiens* selected as the organism. Hits refer to the number of metabolites involved in the pathways, as listed in the 'Metabolites' column. The pathways were deemed statistically significant if they had Hits $\geqslant 2$, a p-value $< 0.05$, and an adjusted p-value obtained using the Holm-Bonferroni test (Holm p) and a False Discovery Rate (FDR) $< 0.05$.

**Table S9. The Out-Of-Bag (OOB) error was calculated using a random forest approach to classify clustering patterns among metabolomic profiles of whole cohort of PD and iPD, within gender.**

| | PD_F | PD_M | OOB error | Total OOB |
|---|---|---|---|---|
| PD_F | 101 | 15 | 0.12 | 0.06 |
| PD_M | 1 | 128 | 0.0007 | |
| | **iPD_F** | **iPD_M** | **OOB error** | **Total OOB** |
| iPD_F | 48 | 8 | 0.14 | 0.07 |
| iPD_M | 8 | 64 | 0.01 | |

The table summarizes the performance of the Random Forest classification models applied to PD patients whole cohort and idiopathic PD patients stratified by sex. For each model, the class error (class.error) and the OOB error are reported, with the latter serving as an internal estimate of the model's accuracy.

**Table S10. Robust Volcano plot results related to Idiopathic PD patients compared to HC with gender stratification**

| Metabolites iPD_M vs HC_M | FC | log2(FC) | raw.pval | -log10(p) |
|---|---|---|---|---|
| L-Glutamic acid | 0.3954 | -1.3386 | 8.14E-12 | 11.089 |
| L-Tryptophan | 2.6266 | 1.3932 | 3.20E-10 | 9.4953 |
| Pyruvic acid | 0.49791 | -1.0061 | 7.60E-07 | 6.1194 |
| Acetic acid | 0.43645 | -1.1961 | 9.35E-06 | 5.0291 |
| L-Methionine | 1.6259 | 0.70124 | 0.000138 | 3.8587 |
| Malonate | 0.59178 | -0.75688 | 0.000241 | 3.6182 |
| Choline | 0.63567 | -0.65365 | 0.000646 | 3.1896 |
| **Metabolites iPD_F vs HC_F** | **FC** | **log2(FC)** | **raw.pval** | **-log10(p)** |
| L-Threonine | 2.7858 | 1.4781 | 2.51E-07 | 6.6002 |
| Choline | 0.55807 | -0.84149 | 4.02E-07 | 6.3962 |
| Acetic acid | 0.49788 | -1.0061 | 4.85E-07 | 6.3145 |

| | | | | |
|---|---|---|---|---|
| 3-Hydroxybutyric acid | 1.5845 | 0.66401 | 0.003375 | 2.4717 |

Results from univariate analysis were presented using a Robust Volcano plot of metabolite concentrations detected in the 1d-CPMG NMR spectrum. The analysis combines the Fold change (FC) test, calculated as the ratio of average concentrations between the iPD_M/HC_M and iPD_F/HC_F clusters—using a threshold of 1.5—and the p-value from the T-Test, which is considered significant if less than 0.05

**Table S11. Pathway enrichment analysis across male and female iPD patients**

| Dysregulated Pathways iPD_M | Hits | Raw p | Holm p | FDR | Metabolites |
|---|---|---|---|---|---|
| Tryptophan Metabolism | 4 | 1.11E-04 | 7.74E-04 | 7.74E-04 | L-Glutamic acid; L-Tryptophan; Formate; L-Alanine. |
| Glutathione Metabolism | 3 | 5.70E-03 | 3.94E-03 | 2.00E-03 | Glycine; L-Glutamic acid; L-Ornithine. |
| Glycine and Serine Metabolism | 11 | 5.37E-03 | 3.65E-03 | 1.25E-03 | Betaine, Creatine, Glycine, L-Glutamic acid, L-Alanine, L-Threonine, L-Serine, L-Ornithine, Pyruvic acid, L-Methionine, L-Arginine. |
| Cysteine Metabolism | 2 | 2.29E-03 | 1.54E-03 | 3.51E-03 | L-Glutamic acid, Pyruvic acid. |
| Malate-Aspartate Shuttle | 2 | 3.54E-03 | 2.33E-03 | 3.51E-03 | Aspartate; L-Glutamic acid. |
| Lysine Degradation | 2 | 3.64E-03 | 2.33E-03 | 3.51E-03 | L-Glutamic acid; Lysine. |
| Alanine Metabolism | 4 | 4.28E-03 | 2.70E-03 | 3.51E-03 | Glycine; L-Glutamic acid; L-Alanine; Pyruvic acid. |
| Nicotinate and Nicotinamide Metabolism | 2 | 4.52E-03 | 2.80E-03 | 3.51E-03 | L-Glutamic acid; L-Glutamine. |
| Glucose-Alanine Cycle | 4 | 8.22E-03 | 5.01E-03 | 5.75E-03 | D-Glucose; L-Glutamic acid; L-Alanine; Pyruvic acid. |
| Tyrosine Metabolism | 4 | 9.58E-03 | 5.75E-03 | 6.10E-03 | Acetoacetic acid; L-Glutamic acid; Tyrosine; L-Phenylalanine. |
| Folate Metabolism | 2 | 4.79E-03 | 2.83E-03 | 2.80E-03 | Formate; Glutamic acid. |
| Histidine Metabolism | 3 | 7.45E-03 | 4.32E-02 | 4.01E-03 | 1-Methylhistidine; L-Glutamic acid; L-Histidine. |
| Phenylalanine and Tyrosine Metabolism | 4 | 1.15E-03 | 6.54E-02 | 5.74E-03 | Aceto acetic; Tyrosine; L-Phenylalanine; L-Glutamic acid. |
| Ammonia Recycling | 8 | 1.31E-03 | 7.35E-02 | 6.13E-03 | L-Glutamic acid; Pyruvic acid; L-Asparagine; Glycine; Serine; Aspartate; L-Histidine, L-Glutamine. |
| **Dysregulated Pathways iPD_F** | **Hits** | **Raw p** | **Holm p** | **FDR** | **Metabolites** |
| Sphingolipid Metabolism | 2 | 2.14E-05 | 1.39E-03 | 2.21E-03 | L-Serine; D-Glucose. |
| Methionine Metabolism | 5 | 2.21E-05 | 1.42E-03 | 2.21E-03 | Betaine; Glycine; L-Serine; Choline; L-Alanine. |
| Phosphatidylethanolamine Biosynthesis | 2 | 2.72E-05 | 1.82E-04 | 4.08E-02 | Choline; L-Serine. |
| Ammonia Recycling | 8 | 1.93E-05 | 4.59E-04 | 4.50E-02 | L-Glutamic acid; Pyruvic acid; L-Asparagine; Glycine; L-Serine; Aspartate; L-Histidine, L-Glutamine |
| Selenoamino Acid Metabolism | 2 | 1.23E-03 | 8.64E-03 | 4.43E-03 | L-Alanine; L-Serine. |

List of enriched pathways resulted from the comparison of male and female iPD patients' serum profiles compared to matched HC. The analysis was performed using the Small Molecule Pathways Database (SMPDB) with *Homo sapiens* selected as the organism. Hits refer to the number of metabolites involved in the pathways, as listed in the 'Metabolites' column. The pathways were deemed statistically significant if they had Hits $\geqslant 2$, a p-value $< 0.05$, and an adjusted p-value obtained using the Holm-Bonferroni test (Holm p) and a False Discovery Rate (FDR) $< 0.05$.

**Table S12. Robust Volcano plot results related to genetic PD patients compared to HC with gender stratification**

| Metabolites gPD_M vs HC_M | FC | log2(FC) | raw.pval | -log10(p) |
|---|---|---|---|---|
| L-Glutamic acid | 0.39083 | -1.3554 | 1.31E-16 | 15.884 |
| L-Tryptophan | 2.8329 | 1.5023 | 4.46E-10 | 9.3505 |
| L-Methionine | 1.8069 | 0.8535 | 1.62E-07 | 6.7915 |
| Acetic acid | 0.41879 | -1.2557 | 2.07E-06 | 5.6849 |
| Choline | 0.61311 | -0.70579 | 2.69E-05 | 4.571 |
| Malonate | 0.6293 | -0.66818 | 0.000114 | 3.9421 |
| Pyruvic acid | 0.55931 | -0.83828 | 0.000201 | 3.6963 |
| **Metabolites gPD_F vs HC_F** | **FC** | **log2(FC)** | **raw.pval** | **-log10(p)** |
| Choline | 0.59969 | -0.73772 | 1.2E-13 | 12.993 |
| Acetic acid | 0.50086 | -0.99753 | 7.99E-13 | 12.097 |
| L-Threonine | 1.8633 | 0.89789 | 1.37E-09 | 8.8631 |
| 3-Hydroxybutyric acid | 1.7519 | 0.80892 | 1.67E-06 | 5.7765 |
| Pyruvic acid | 0.65783 | -0.60421 | 3.31E-05 | 4.4796 |
| L-Methionine | 1.5693 | 0.65017 | 3.54E-05 | 4.4513 |

Results from univariate analysis were presented using a Robust Volcano plot of metabolite concentrations detected in the 1d-CPMG NMR spectrum. The analysis combines the Fold change (FC) test, calculated as the ratio of average concentrations between the gPD_M/HC_M and gPD_F/HC_F clusters—using a threshold of 1.5—and the p-value from the T-Test, which is considered significant if less than 0.05.

**Table S13. Pathway enrichment analysis across male and female gPD patients**

| Dysregulated Pathways gPD_M | Hits | Raw p | Holm p | FDR | Metabolites |
|---|---|---|---|---|---|
| Cysteine Metabolism | 2 | 1.41E-08 | 9.70E-07 | 4.92E-07 | L-Glutamic acid, Pyruvic acid. |
| Malate-Aspartate Shuttle | 2 | 2.96E-08 | 2.02E-06 | 6.91E-07 | Aspartate; L-Glutamic acid. |
| Folate Metabolism | 2 | 6.78E-08 | 4.54E-07 | 1.19E-06 | Formate; L-Glutamic acid. |
| Amino Sugar Metabolism | 4 | 1.91E-07 | 1.26E-05 | 2.67E-06 | Acetic acid; L-Glutamic acid; Pyruvic acid; L-Glutamine |
| Histidine Metabolism | 3 | 5.32E-07 | 3.46E-05 | 6.21E-06 | 1-Methylhistidine; L-Glutamic acid; L-Histidine. |
| Beta-Alanine Metabolism | 3 | 1.59E-06 | 1.02E-04 | 1.59E-05 | L-Glutamic acid; Aspartate; L-Histidine. |
| Glucose-Alanine Cycle | 4 | 2.97E-06 | 1.87E-04 | 2.60E-05 | D-Glucose; L-Glutamic acid; L-Alanine; Pyruvic acid. |
| Tryptophan Metabolism | 4 | 3.90E-06 | 2.42E-05 | 3.03E-05 | Formate; L-Glutamic acid; L-Alanine; L-Tryptophan. |
| Lysine Degradation | 2 | 7.92E-06 | 4.83E-04 | 5.54E-05 | L-Glutamic acid; L-Lysine. |
| Nicotinate and Nicotinamide Metabolism | 2 | 1.36E-06 | 8.17E-05 | 8.67E-05 | L-Glutamic acid; L-Glutamine. |
| Alanine Metabolism | 4 | 3.29E-05 | 1.94E-03 | 1.92E-04 | Glycine; L-Glutamic acid; L-Alanine; Pyruvic acid. |
| Glutathione Metabolism | 3 | 8.36E-05 | 4.85E-03 | 4.50E-04 | Glycine; L-Glutamic acid; L-Ornithine |
| Aspartate Metabolism | 6 | 3.26E-04 | 1.86e-06 | 1.63E-03 | L-Glutamic acid; Acetic acid; Asparagine; Aspartate; L-Glutamine; L-Arginine. |
| Propanoate Metabolism | 3 | 3.57E-04 | 2.00E-02 | 1.66E-03 | 2-Hydroxybutyric acid; L-Glutamic acid; L-Valine. |
| **Dysregulated Pathways gPD_F** | **Hits** | **Raw p** | **Holm p** | **FDR** | **Metabolites** |
| Fatty Acid Biosynthesis | 3 | 1.10E-12 | 7.60E-11 | 3.86E-11 | Acetic acid, Acetoacetic acid, 3-Hydroxybutyrate |
| Pyruvic acid Metabolism | 3 | 6.44E-12 | 4.25E-10 | 8.49E-11 | Acetic acid; Lactic acid; Pyruvic acid |
| Amino Sugar Metabolism | 4 | 7.28E-12 | 4.73E-10 | 8.49E-11 | Acetic acid; Pyruvic acid; L-Glutamine; L-Glutamic acid |
| Betaine Metabolism | 3 | 2.62E-10 | 1.68E-08 | 2.62E-09 | Betaine; Choline; L-Methionine |
| Methionine Metabolism | 5 | 2.61E-09 | 1.65E-07 | 2.29E-08 | Betaine; Glycine; L-Serine; L-Methionine;L-Choline |
| Phosphatidylethanolamine Biosynthesis | 2 | 1.88E-08 | 1.16E-06 | 1.46E-07 | L-Choline; L-Serine |

**Checklist**

**1. Reporting Guidelines**
-STROBE Compliance: This study was designed and reported following the Strengthening the Reporting of Observational Studies in Epidemiology (STROBE) guidelines for case-control studies. **Pag. 23 lines 15-16**

-Metabolomics Reporting: Metabolomic data acquisition and processing comply with the Metabolomics Standards Initiative (MSI) guidelines. **Pag. 23 lines 15-16**

**2. Ethical Compliance & Registration**
**-**Institutional Review Board (IRB): Approved by the IRB of IRCCS Neuromed, Italy (Protocols: N°9/2015, N°19/2020, N°4/2023). **Pag. 23 lines 8-10**

-ClinicalTrials.gov Registration: The study is registered under identifiers NCT02403765, NCT04620980, and NCT05721911. **Pag. 23 lines 9-10**

-Declaration of Helsinki: All clinical investigations were conducted according to the principles of the Declaration of Helsinki. **Pag. 23 lines 11-12**

-Informed Consent: Written informed consent was obtained from all study participants. **Pag. 23 line 12**

**3. Data Stratification & Quality Control**
- Genetic Stratification: Patients were stratified into two distinct groups:
iPD: Idiopathic (no variants in PD genes).
gPD: Genetic (pathogenic mutations in LRRK2, GBA1, TMEM175, PARK2, PINK1, PARK7). **Pag. 5 lines 3-8**

- Sex-Matching: Healthy controls (HC) were sex-matched to the PD cohort to minimize confounding bias in metabolomic analysis. **Pag. 23 Lines 1-3**

- Confounder Adjustment: Statistical analyses were adjusted for age, disease duration, and L-DOPA Equivalent Daily Dose (LEDD). **Pag. 19 lines 18-24**

**4. Technical Validation**
- NMR Quantitation: Metabolite identification and quantification were performed using Chenomx NMR Suite and confirmed with Bayesil software. **Pag. 25 lines 21-25; pag 26 lines 1,2.**

**5. Data Availability & Reproducibility**
- Software used: MetaboAnalyst 6.0 (Metabolomics) **Pag.26 lines 20-21;** Chenomx (NMR) **Pag. 25 line 21,** Small Molecules Pathways Database (SMPDB) **Pag. 27 Lines 13-14**; Permutational Multivariate Analysis of Variance (PERMANOVA) test **Pag. 26 Lines 17-19.**

- Public Databases: NA

**1. Study Design & Setting (STROBE Items 4 & 5)**
- Design Type: Defined as a case-control observational study. **Pag.22 line 3**

- Study Periods: Two recruitment windows specified (June 2015–Dec 2017 and June 2021–Dec 2023) **Pag. 22 lines 9-13.**

- Location: Parkinson Centre of the IRCCS INM Neuromed, Italy **Pag 22. Lines 9-13.**

**2. Participant Selection & Eligibility (STROBE Item 6)**
-Case Definition: PD diagnosis based on ≥2 cardinal motor signs (tremor, bradykinesia, rigidity) and positive response to L-DOPA. **Pag. 22 lines 16-18**

-Control Definition: Healthy subjects (HC) negative for PD gene mutations, matched for sex with the PD cohort. **Pag.23 lines 3-5.**

-Age Threshold: Inclusion limited to individuals aged ≥40 years to maintain cohort relevance. **Pag.23 line 3**

- Exclusion Criteria: Explicitly listed (pre-existing psychiatric conditions, other neurodegenerative diseases like MS or ALS, dementia, depression, and use of specific psychotropic medications). **Pag. 22 lines 18-22**

**3. Data Sources & Clinical Assessment (STROBE Item 8)**
-Clinical Scale: MDS-UPDRS Part III used for motor symptom severity (assessed during the "ON" period). **Pag. 22 lines 23-24; pag.23 lines 1-2.**

-Ancestry: Confirmed European ancestry for all participants. **Pag. 22 line 10**

-Biobank Origin: Subjects selected from the IRCCS Neuromed/IGB-CNR biobank. **Pag.22 line 9**

**4. Genetic Stratification Framework**
- Sequencing Method: Whole Exome Sequencing (WES) data analyzed for the presence of mutations/variants in PD genes. **Pag.21 lines 4-6.**

- Group Classification:
iPD (Idiopathic): No mutations in PD genes. **Pag. 5 lines 4-5.**
gPD (Genetic): Carrying known pathogenic mutations (e.g., LRRK2 G2019S, GBA1). **Pag. 5 lines 6-8.**

**5. Laboratory & Analytical Protocols**
- Serum Handling: Standardized 6-hour fasting collection, 30-min clotting, and -80°C storage. **Pag. 24 lines 20-24.**

- NMR Parameters: Bruker DRX600 MHz spectrometer **Pag.25 lines 11-13**, CPMG pulse sequence **Pag.25 line 19**,

- Standardization: Use of anonymized codes and internal reference signals (TSP) **Pag. 25 lines 9-11.**

**6. Statistical & Confounding Control (STROBE Items 10 & 12)**
- Sample Size: Acknowledged as determined by biobank availability (no formal a priori calculation). **Pag. 23 lines 19-20**

- Normality Testing: Metabolite concentrations were log-transformed prior to analysis to improve normality assumptions; robust (HC3) standard errors were also applied to account for deviations from normality and heteroscedasticity. **Pag. 27 Line 26; pag. 28 lines 1-3.**

- Confounder Adjustment: Models were adjusted for relevant covariates, including age, sex, disease duration, and LEDD, with sex also included as an interaction term where appropriate. **Pag. 19 lines 18-24**

- Multivariate Analysis: Multivariate analyses included PCA (validated by PERMANOVA), PLS-DA with cross-validation (R2, Q2), and Random Forest classification with OOB error estimation. **Pag. 6 lines 5-11.**

- Multiple Testing: Multiple comparisons were controlled using the Benjamini–Hochberg FDR correction across metabolite-level and enrichment analyses, with FDR $< 0.05$ considered significant. **Pag.28 lines 20-23.**
